# Comparison of seroprevalence of SARS-CoV-2 infections with cumulative and imputed COVID-19 cases: systematic review

**DOI:** 10.1101/2020.07.13.20153163

**Authors:** Oyungerel Byambasuren, Claudia C Dobler, Katy Bell, Diana Patricia Rojas, Justin Clark, Mary-Louise McLaws, Paul Glasziou

**Affiliations:** Institute for Evidence-Based Healthcare, Bond University; School of Public Health, University of Sydney; College of Public Health, Medical and Veterinary Sciences, Division of Tropical Health & Medicine, James Cook University; School of Public Health and Community Medicine, UNSW Sydney

**Author notes:** **Corresponding author**: Oyungerel Byambasuren, Institute for Evidence-Based Healthcare Bond University, 14 University Dr, Robina QLD 4226 Australia.

**Keywords:** seroprevalence, COVID-19, SARS-CoV-2, systematic review, herd immunity

## Abstract

**Background:** Accurate seroprevalence estimates of SARS-CoV-2 in different populations could clarify the extent to which current testing strategies are identifying all active infection, and hence the true magnitude and spread of the infection. Our primary objective was to identify valid seroprevalence studies of SARS-CoV-2 infection and compare their estimates with the reported, and imputed, COVID-19 case rates within the same population at the same time point.

**Methods:** We searched PubMed, Embase, the Cochrane COVID-19 trials, and Europe-PMC for published studies and pre-prints that reported anti-SARS-CoV-2 IgG, IgM and/or IgA antibodies for serosurveys of the general community from 1 Jan to 12 Aug 2020.

**Results:** Of the 2199 studies identified, 170 were assessed for full text and 17 studies representing 15 regions and 118,297 subjects were includable. The seroprevalence proportions in 8 studies ranged between 1%-10%, with 5 studies under 1%, and 4 over 10% - from the notably hard-hit regions of Gangelt, Germany; Northwest Iran; Buenos Aires, Argentina; and Stockholm, Sweden. For seropositive cases who were not previously identified as COVID-19 cases, the majority had prior COVID-like symptoms. The estimated seroprevalences ranged from 0.56-717 times greater than the number of reported cumulative cases – half of the studies reported greater than 10 times more SARS-CoV-2 infections than the cumulative number of cases.

**Conclusions:** The findings show SARS-CoV-2 seroprevalence is well below “herd immunity” in all countries studied. The estimated number of infections, however, were much greater than the number of reported cases and deaths in almost all locations. The majority of seropositive people reported prior COVID-like symptoms, suggesting that undertesting of symptomatic people may be causing a substantial under-ascertainment of SARS-CoV-2 infections.

**Key messages:**

- Systematic assessment of 17-country data show SARS-CoV-2 seroprevalence is mostly less than 10% - levels well below “herd immunity”.
- High symptom rates in seropositive cases suggest undertesting of symptomatic people and could explain gaps between seroprevalence rates and reported cases.
- The estimated number of infections for majority of the studies ranged from 2-717 times greater than the number of reported cases in that region and up to 13 times greater than the cases imputed from number of reported deaths.

## Introduction

Globally, over one hundred million coronavirus disease (COVID-19) cases have been reported to World Health Organization as of 15 February 2021.[1] However, seroprevalence estimates based on immune response (serum antibodies) to SARS-CoV-2 rather than reverse transcriptase polymerase chain reaction (RT-PCR) testing [2], may provide a more accurate reflection of the true extent of SARS-CoV-2 infection among a population as many people may have not been tested when they had active infection.

Valid seroprevalence estimates for a population rely on two major factors: (i) a representative population sample and (ii) accurate antibody testing. For example, testing should not be biased by including predominantly symptomatic people or those exposed to a person with COVID-19.[3] Inappropriate sampling will bias the estimated seroprevalence, the infection fatality rate, and the effective reproductive number (Rt).[4]

Systematic Reviews of the diagnostic accuracy SARS-CoV-2 antibodies have found concerns about bias and applicability in the available studies. The sensitivity of most antibody-tests, which measure immunoglobulin (Ig) M, IgG, and occasionally IgA antibodies against SARS-CoV-2, appears to be low in the first week after onset of symptoms and increases up to maximum value in the third week; data beyond three weeks are scarce.[5-7] Specificity of the antibody tests has been estimated to exceed 98% for most tests; however, this may still result in poor positive predictive values and high false positive rates in low prevalence settings.[6] Some evidence suggests that in infected asymptomatic people, a reduction of serum antibodies is already observed during the early convalescent phase.[8]

We aimed to identify all studies that reported seroprevalence estimates for SARS-CoV-2 infection using a representative sample of the target population, and to compare to these seroprevalence estimate with the cumulative incidence of confirmed COVID-19 cases, and imputed case rates from the death rates, to establish the likely true extent of the infection among a population.

## Methods

We conducted a systematic review using enhanced processes with initial report completed within two weeks, using daily short team meetings to review the progress, plan next actions, and solve discrepancies and other obstacles.[9] We also used locally developed open access automation tools and programs such as the Polyglot Search Translator, SearchRefiner, and the SRA Helper to design, refine and convert our search strategy for all the databases we searched and to speed up the screening process. We searched PubMed, Embase, Cochrane COVID-19 trials for published studies, and Europe PMC for pre-prints from 1 January to 12 August 2020. A search string composed of Medical Subject Headings (MeSH) terms and words was developed in PubMed and was translated to be run in other databases[10] (see Supporting information S1 file). We also conducted forward and backward citation searches of the included studies in the Scopus citation database. No restrictions on language were imposed. Review protocol was not registered.

### Inclusion Criteria

We included seroprevalence studies which attempted complete or random sample of the population with more than 25% response rate to assess overall seroprevalence in general community. We included seroprevalence testing that tested for anti-SARS-CoV-2 IgG, IgM, and IgA antibodies in combination or separately.

We excluded studies with high risk of bias in sampling, i.e. the study sample was likely not representative of the target population such as health care workers, blood donors, or dialysis patients; government reports without sufficient details to evaluate risk of bias; modelling or simulation studies even if they used real data (but sources of real data were checked for possible inclusion); lack of information about the antibody test(s) used to determine seroprevalence; and editorial or historical accounts without sufficient data to calculate the primary outcome (e.g. insufficient details to allow identification of cumulative reported cases in the population detected using RT-PCR). A list of excluded studies can be found in Supplement 2 with reasons for exclusion.

### Outcomes

Our primary outcomes were (1) the comparison of the seroprevalence based on antibody testing in the study sample with the cumulative reported case incidence of people tested positive for SARS-CoV-2 by RT-PCR in the same sample or in the target population cross-checked by a cumulative incidence estimated from the cumulative COVID-19-specific mortality two weeks after the seroprevalence and assuming a case-fatality rate of 1% [11]; and (2) frequency of COVID-like symptoms among the study population prior to serological testing and odds of testing positive with prominent COVID-related symptoms where data available.

### Study selection and screening

Two authors (OB and CCD) independently screened titles, abstracts, and full texts according to inclusion criteria. All discrepancies were resolved via group discussion with the other authors. Reasons for exclusion were documented for all full text articles deemed ineligible (supporting information S2 Table) - see PRISMA diagram (Figure 1).

**Figure 1.**
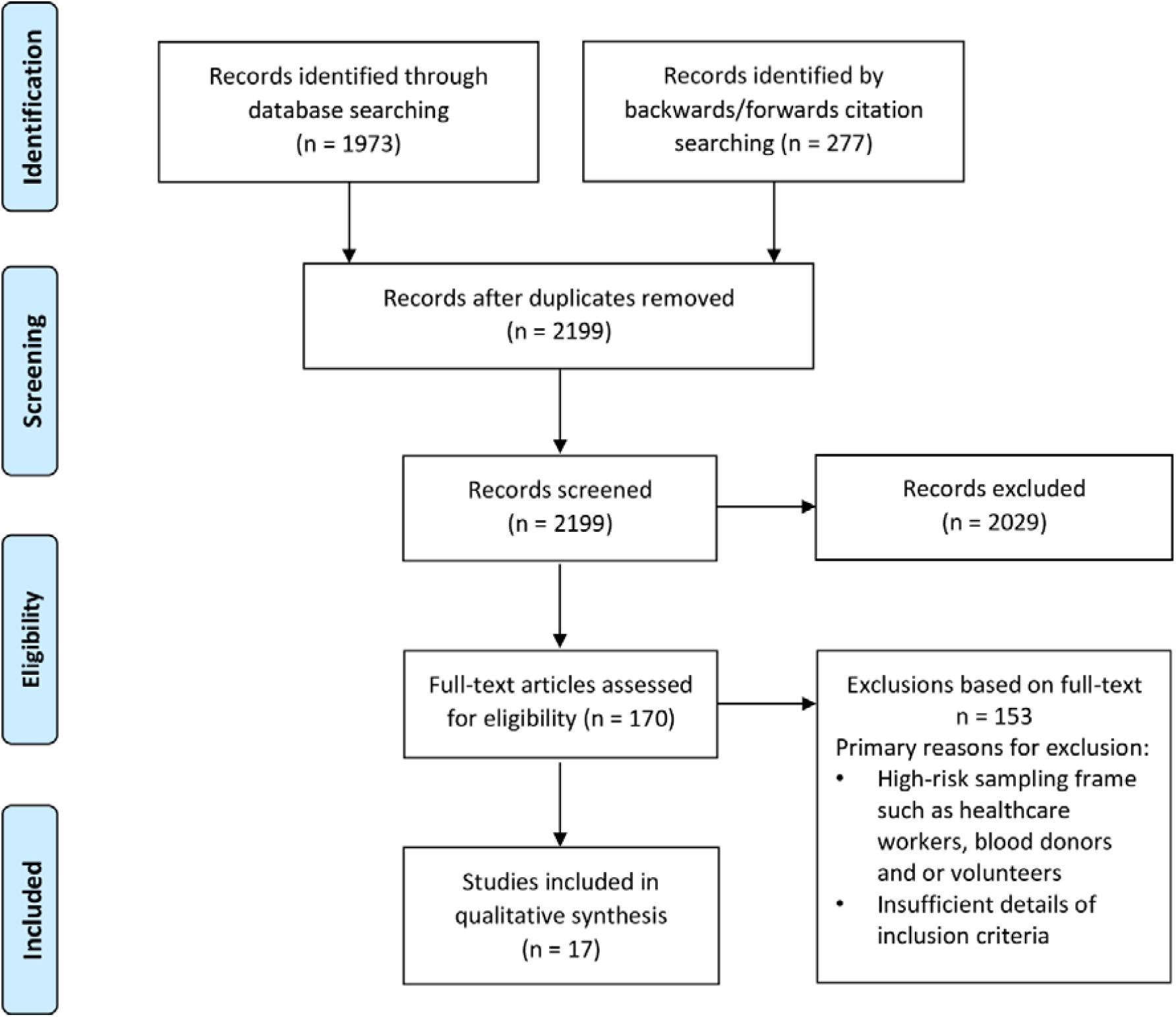
Screening and selection of articles for the review.

### Data extraction

Five authors (OB, CCD, KB, PG, DPR) extracted the following information from each study and from related external sources:

- Participants: sampling frame, sample size, age, sex, setting, previous exposure or testing for COVID-19
- Methods: study authors, country or region of the study, publication type, types of tests used, date of seroprevalence sampling (to enable identification of separately reported cumulative incidence rate in the sampling frame at around the same time as seroprevalence study).
- Outcomes: study seroprevalence (point estimate and confidence interval), adjusted seroprevalence (point estimate for the population adjusted for study design and test accuracy), and cumulative COVID-19 cases in the study sample.
- Other information: when not provided in the study, we looked for publicly available data on the cumulative incidence of COVID-19 and COVID-19 specific mortality in the study population as close to the time of the study as possible.

### Risk of bias assessment

We used a combination of risk of bias tools for prevalence studies[12] and diagnostic accuracy[13] and adapted the key signaling questions on sampling frame, ascertainment of immune status, acceptability of methods and tests, and appropriateness of testing and sample collection timeframe, as shown in supporting information S3 in full.

### Data synthesis

We used absolute numbers and proportions for the primary outcome. As only studies deemed to be of sufficient quality after critical appraisal were included in the analysis, no sensitivity analysis of high versus low quality studies was undertaken. We did not pool the estimates due to heterogeneity of populations and study methods.

## Results

We screened titles and abstracts of 2,199 articles and the full text of 170 articles for potential inclusion (Figure 1). The major reason for exclusion was high risk of bias in the selection of participants (Full list of excluded studies in Supplement 2). Seventeen articles – 4 preprints, 11 published studies, and 2 government reports– from 15 countries (Argentina, Brazil, Spain, Hungary, Germany, Luxembourg, Switzerland, Denmark, Sweden, Finland, Iceland, the United States of America (USA), the Channel Islands, Iran, and Japan) that tested a combined total of 118,297 participants met eligibility criteria.[14-30] (Table 1.)

**Table 1.** Characteristics of included studies (n=17)

| **Study region, country, author, publication status** | **Study population (sampling frame)** | **Sample size, mean age, sex, study dates** | **Type of serologic test and their Sensitivity and Specificity** |
| --- | --- | --- | --- |
| **Spanish national sero-epidemiological survey**<br>Pollán et al [20]<br>Published | Randomly selected population of Spain from census data | n=61,075<br>mean age 44 years<br>52% female<br>27 April - 11 May | IgG and IgM: Orient Gene IgM/IgG, Zhejiang Orient Gene Biotech |
| **Brazilian nationwide survey**<br>Hallal et al [16]<br>Preprint | Random samples of 133 large sentinel cities from all 26 states and the Federal District in Brazil | n=24,995<br>mean age 43<br>58% female<br>14 -21 May | IgG and IgM: WONDFO 459 SARS-CoV-2 Antibody Test (Wondfo Biotech Co., Guangzhou, China) |
| **Hungary**<br>Merkely et al [17]<br>Published | Random sampling of representative Hungarian population over 14 years of age. | n=10,474<br>mean age 49 years<br>53.6% female<br>1-16 May | IgG: SARS-CoV2 IgG Reagent Kit, Abbott Laboratories, Irving, TX, USA |
| **Luxembourg**<br>Snoeck et al [24]<br>Preprint | Random sample of Luxembourg population over age of 18 (n=514,921) | n=1820<br>mean age 47 years<br>51% female<br>16 April - 5 May | IgG and IgA: CE-labelled ELISA kits most recent versions from Euroimmun. |
| **Rio Grande do Sul, Brazil**<br>Silviera et al [23]<br>Published | Random sample of population in Rio Grande do Sul state (population 11.3mln) | n=4500<br>mean age 48 years<br>59% female<br>9-11 May | IgG and IgM: WONDFO SARS-CoV-2 Antibody Test (Wondfo Biotech Co., Guangzhou, China) |
| **Faroe Island, Denmark**<br>Petersen et al [19]<br>Published | Randomly selected population of the island (population 52,154), | n=1075<br>Mean age 42 years<br>50% female<br>27 April-1 May | IgG and IgM: SARS-CoV-2 Ab ELISA kit (Beijing Wantai Biologic Pharmacy Enterprise) |
| **LA county, USA**<br>Sood et al [25]<br>Published | Random sample of LA county population | n=863<br>mean age 44 years<br>60% female<br>10-14 April | IgG and IgM: Lateral Flow Immunoassay test (Premier Biotech) |
| **Jersey Island**<br>The Channel Islands [29]<br>Report | Random sample of adult resident population of Island of Jersey living in private households | n=855<br>mean age 48 years<br>53% female<br>29 April - 5 May | IgG and IgM: Lateral Flow Immunoassay (Healgen COVID-19 IgG/IgM) |
| **Guilan, Iran**<br>Shakiba et al [22]<br>Published | Random sample of population of Guilan province, Iran (population 2,354,848) | n=528<br>mean age 35 years<br>51% female<br>April | IgG and IgM: VivaDiag COVID-19 IgM/IgG from VivaChek |
| **Reykjavik, Iceland**<br>Gudbjartsson et al [15]<br>Published | Population of greater Reykjavik area who had not been tested with PCR or had been tested and negative | n=4843<br>Mean age 48 years<br>38% female<br>27 April – 5 June | pan-Ig: IgM, IgG, & IgA against nucleoprotein (N) (Roche); the receptor binding domain (Wantai); IgM & IgG against N (EDI/Eagle); and IgG & IgA against the spike protein (Euroimmun). |
| **Geneva, Switzerland**<br>Stringhini et al [27]<br>Published | Random sample of Bus Santé study participants, canton of Geneva | n=1956<br>mean age 44 years<br>53% female<br>20 Apr-10 May | IgG: commercially available ELISA for IgG (Euroimmun AG, Lübeck, Germany) |
| **Stockholm, Sweden**<br>Roxhed et al [21]<br>Preprint | Random household sample of adults (20-74 years) in Stockholm | n=1097<br>Mean age 47 years<br>55% female<br>April-May | IgG: commercially available ELISA for IgG against S1 and N proteins |
| **Five university hospital districts, Finland**<br>Finnish Institute for Health and Welfare Report [28] | Random sampling of adult population from 5 hospital districts in southern Finland since 1 June | n=1056<br>Age range 18-69 years<br>1 June – 6 Sep | IgG: against nucleoprotein and spike glycoprotein S1 and S2, the antigens manufactured by The Native Antigen Company |
| **Gangelt, Germany**<br>Streeck et al [26]<br>Published | Random sample of population of Gangelt, Germany (n=12,597) from civil register | n=919<br>mean age 53 years<br>51% female<br>31 Mar- 6 Apr | IgG and IgA: ELISA on the EUROIMMUN Analyzer I platform (most recent CE version for IgG ELISA as of April 2020) |
| **Barrio Mugica, Buenos Aires, Argentina**<br>Figar et al [14]<br>Preprint | Random sample of residents over 14 years of age, Barrio Mugica slum (n= 40,000), Buenos Aires city | n=873<br>median age 38 years<br>57% female<br>*10-26 June* | IgG: COVIDAR IgG ELISA (Laboratorio Lemos SRL, Buenos Aires, Argentina) |
| **Utsunomiya, Japan**<br>Nawa et al [18]<br>Published | Random selection of residents in Utsunomiya City in Tochigi Prefecture, Greater Tokyo, Japan | n=742<br>Mean age 44 years<br>52.6% female<br>*14 June-5 July* | IgG: SARS-CoV2 IgG chemiluminescence assay from Shenzhen YHLO Biotech Co., Ltd., Shenzhen, China |
| **Neustadt-am-Rennsteig, Germany**<br>Weis et al [30]<br>Published | Whole population of Neustadt-am-Rennsteig village, Germany (population 883) | N=626<br>Mean age 60 years<br>53% female<br>*12-22 May* | IgG: two ELISA (Epitope Diagnostics Inc., San Diego, USA, Euroimmun, Lübeck, Germany) and four chemiluminescence assays (DiaSorin, Saluggia, Italy, Snibe Co., Ltd., Shenzhen, China, Abbott, Chicago, USA, and Roche, Basel Switzerland) |

Four studies provide national level data [16, 17, 20, 24], five studies report a province, county or self-governing area level data [19, 22, 23, 26, 29], and the rest provide a city, town, village or district level data. Seven studies tested participants over the age of 14 years [14, 17, 21, 24, 25, 28, 29] and ten tested population of all ages - the proportion of children and young people (0-19 years) ranged from 7% to 26% and the proportion of participants aged over 60 years ranged from 7% to 37%. Eight studies tested for anti-SARS-CoV-2 IgG only or IgG and IgA, the rest tested for IgG and IgM. (Table 1) Only five of the studies also collected nasopharyngeal swabs for RT-PCR testing at the same time as serologic testing.[15, 17, 22-24] Information on the serological test sensitivity and specificity is provided in S4 table.

### Seroprevalence

The seroprevalences ranged considerably (Table 2 and Figure 2): eight studies reported seroprevalence between 1%-10%; five studies had estimates under 1% [15, 17-19, 23] and four studies had estimates over 10%.[14, 21, 22, 26] The unadjusted and adjusted seroprevalence estimates in the included studies ranged from 0.22% in Rio Grande do Sul state in Brazil [23] to 53% in the Barrio Mugica slum of Buenos Aires, Argentina [14].

**Table 2.**
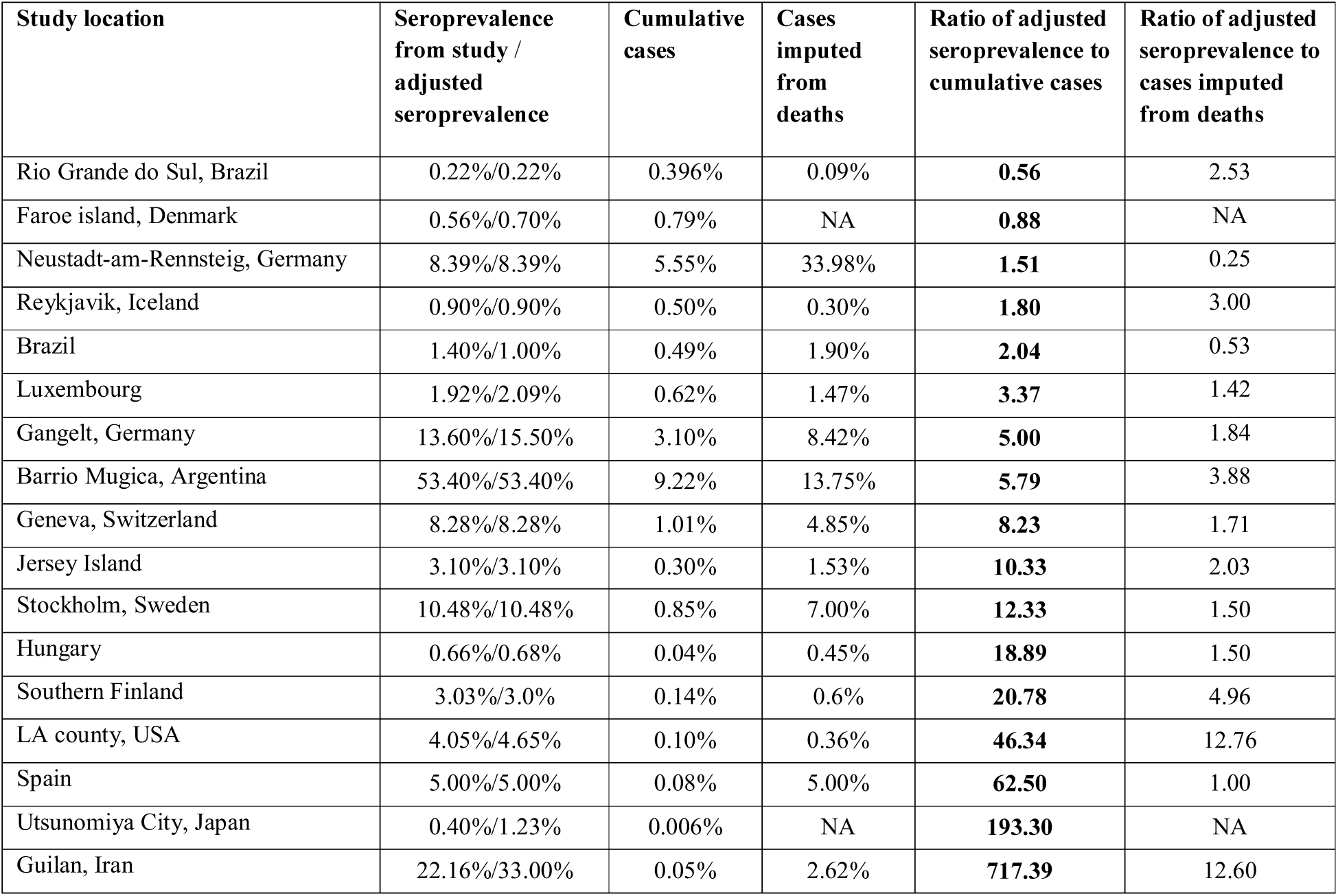
Estimated cumulative incidence of infections based on seroprevalence estimates and compar ison with the number of reported cases and imputed cases from death rate.

**Figure 2.**
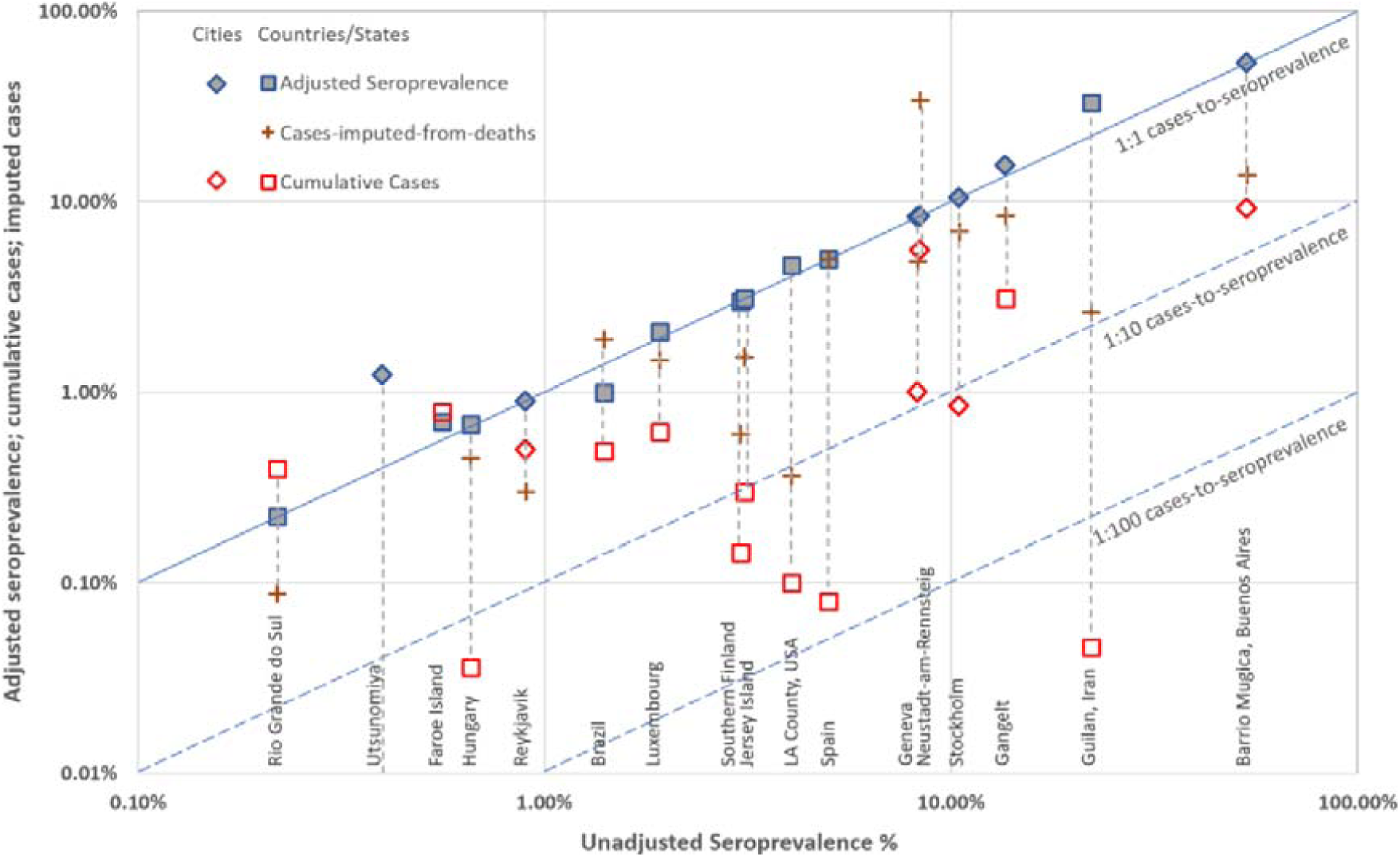
Log-log plot of study seroprevalence (x-axis) vs two cumulative case estimator s for each study. Diagonal lines indicate rates equal to ser opr evalence (solid) or 1/10 seroprevalence (dashed).

The cumulative case incidence in the study population (based on RT-PCR testing) was reported in five studies [15, 17, 22-24]. For the other studies we identified cumulative case incidence data from publicly available online reports. For some studies the two types of estimate were similar (e.g. Faroe island, Denmark), but for others the seroprevalence estimate was substantially higher than the cumulative case estimate (e.g. in Guilan, Iran). Further details on the study adjustment details and sources for cumulative incidence data are provided in S4 Table.

The cumulative incidence rates at the regional levels (red squares and diamonds) ranged from 0.006% in Utsunomiya, Tokyo [18] to 9.22% in Barrio Mugica slum of Buenos Aires, Argentina [14]. The calculated cumulative case incidence for regions imputed from reported COVID-19 deaths (assuming true CFR of 1%, brown crosses) ranged from 0.09% in Rio Grande do Sul, Brazil [23] to 33.98% in Neustadt-am-Rennsteig, Germany[30]. The data collection timeframes of the included studies are shown in S5 Figure in relation to the rolling 7-day average of confirmed cases in each country.

The relationship between all the outcome estimates for each study/region on the log scale are shown in Figure 2. The upper diagonal (identity) line indicates estimates that are equal to the study seroprevalence estimate, and the lower diagonal line indicates estimates that are 1/10 or 1/100 that of the study seroprevalence estimate. In general, cases imputed from reported deaths are next closest to the seroprevalence estimates, although there is considerable variation in how close: imputed cases for Spain[20] matched the seroprevalence almost exactly, while those for Guilan, Iran[22] were around 1/10 of the seroprevalence. Next closest were the study cumulative case estimates, where differences in test accuracy of antibody vs RT-PCT tests may explain most of the within study differences. The estimates that differed the most from those of the study seroprevalence (furthest away from the identity line) were the reported regional case estimates, with several falling below the 1/100 seroprevalence line, some notably so (Guilan, Iran)[22].

### Ratio of seroprevalence to cumulative cases

Table 2 compares estimates of seroprevalence estimates to the cumulative reported cases. For two studies - Rio Grande do Sul, Brazil, and the Faroe Islands, the seroprevalence was less than cumulative cases, but numbers were small. For seven other studies the ratio was less than 10. The highest ratio was in Guilan, Iran, where the estimation of infections was 717 times greater than the reported cases as of April 2020. Two studies did not report any COVID-19 related deaths among the participants so we could not impute case estimates for these studies. [17, 18] For those studies we could impute the cumulative cases from deaths, the ratios were generally much closer to 1, three being less than 1, and only two over 10.

### Symptoms

Typical COVID-like symptoms prior to serologic testing [31] could help assess possible untested or undetected cases. Nine of the 17 studies provided data on prior symptoms and measures varied (Table 3). Between 17% and 83% of the sero-positive participants in six studies reported having typical COVID-like symptoms in the 2 weeks to 3 months prior to the serologic testing. Prevalence of COVID-like symptoms were significantly more common among sero-positive participants compared to the sero-negative participants. Positive serologic testing was 1.5 to 8.1 times more likely in people who had had any acute respiratory infection (ARI) symptoms; for the individual symptoms this ranged from 2-fold (fever) to 46- fold (loss of smell and taste). Three studies also reported prevalence of other non-specific symptoms such as headache, chest pain, skin rash, nausea, and fatigue among the participants. [17, 24, 30]

**Table 3:**
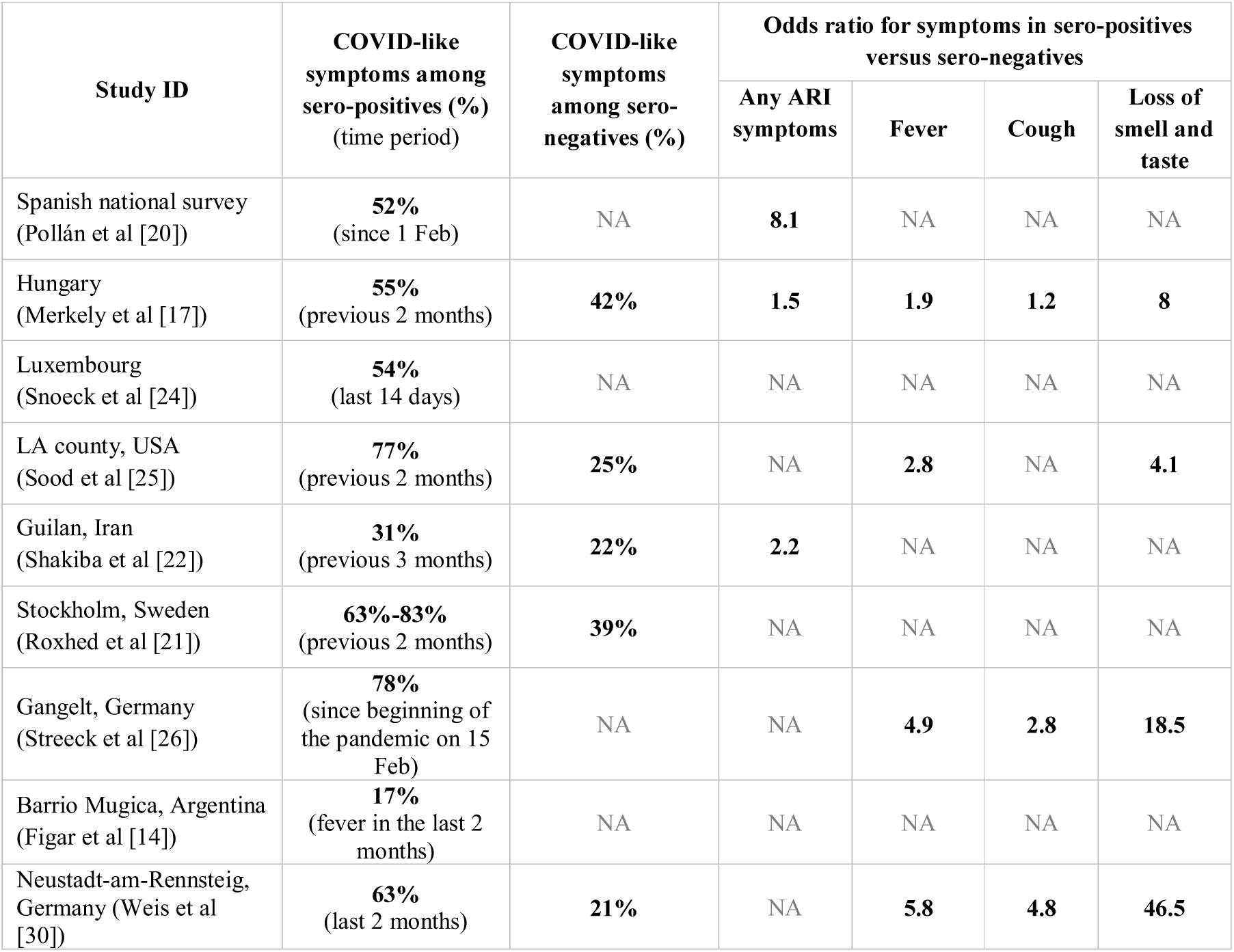
Frequency of COVID-like or respiratory symptoms.

### Risk of Bias of included studies

Table 4 summarizes the overall risk of bias assessment of the 17 included studies (see Supplement 3). Most studies had low risk of bias for the sampling frame as they recruited participants randomly from the general population (Domain 1). Majority of the studies reported response rate over 50%. Five studies reported response rate in lower 30% or unclear (Domain 2). Domain 3 assessed the potential to over- or underestimate the seroprevalence based on the diagnostic accuracy of the individual antibody tests used in each study. Although each study provided specificity and sensitivity for the tests based on internal or external (manufacturer) validation, it was difficult to confidently evaluate the impact on the study results without a single-source validation that would enable unbiased comparison. All studies but one used the same test and type of test specimen in all study participants (Domain 4). The Spanish national serosurvey did not venipuncture children and used only the rapid test (finger prick blood sample) and lab test in adults. We evaluated the appropriateness of the timing of testing as low risk of bias as all studies reported the dates of sample collection and testing as occurring after their local “pandemic wave” had passed. (Domain 5)

**Table 4.** Risk of bias in 14 included studies. Green smiley face denotes low risk of bias; yellow straight face – moderate or unclear risk; and red sad face - high risk of bias.

| Included studies | Risk of bias assessment questions | 1. Was the sampling frame a true or close representation of the target population? | 3. Is the diagnostic test used likely to correctly classify all past infections in the target (at risk) population? | 3. Is the diagnostic test used likely to correctly classify all past infections in the target (at risk) population? | 4. Was the same diagnostic test used for all subjects? | 5. Was the period of testing appropriate? |
| --- | --- | --- | --- | --- | --- | --- |
| Spanish national sero-survey |  | 😊 | 😊 | 😐 | 😐 | 😊 |
| Brazilian nationwide survey |  | 😊 | 😊 | 😐 | 😊 | 😊 |
| Hungary |  | 😊 | 😊 | 😐 | 😊 | 😊 |
| Luxembourg |  | 😊 | 😊 | 😐 | 😊 | 😊 |
| Rio Grande do Sul, Brazil |  | 😊 | 😊 | 😐 | 😊 | 😊 |
| Faroe island, Denmark |  | 😊 | 😊 | 😐 | 😊 | 😊 |
| LA county, USA |  | 😊 | 😊 | 😐 | 😊 | 😊 |
| Jersey Island, Channel Islands |  | 😊 | 😊 | 😐 | 😊 | 😊 |
| Guilan, Iran |  | 😊 | 😐 | 😐 | 😊 | 😊 |
| Reykjavik, Iceland |  | 😊 | 😐 | 😐 | 😊 | 😊 |
| Geneva, Switzerland |  | 😊 | 😐 | 😐 | 😊 | 😊 |
| Stockholm, Sweden |  | 😊 | 😊 | 😐 | 😊 | 😊 |
| Five uni hospital districts, Finland |  | 😊 | 😊 | 😐 | 😊 | 😊 |
| Gangelt, Germany |  | 😊 | 😊 | 😐 | 😊 | 😊 |
| Barrio Mugica, Argentina |  | 😊 | 😐 | 😐 | 😊 | 😊 |
| Utsunomiya, Japan |  | 😊 | 😐 | 😐 | 😊 | 😊 |
| Neustadt-am-Rennsteig, Germany |  | 😊 | 😊 | 😐 | 😊 | 😊 |

## Discussion

The seroprevalence rates in eight studies ranged between 1%-10%, with 5 studies under 1%,and 4 studies over 10% - notably hard-hit regions of Gangelt, Germany, Northwest Iran, the Barrio Mugica slum of Buenos Aires, Argentina, and Stockholm, Sweden. For all but two studies, the seroprevalence estimate was higher than the cumulative reported case incidence, by a factor between 1.5 to 717 times higher. However, the seroprevalence estimates were generally much closer to the cumulative incidence imputed from deaths. Finally, we noted that many of the seropositive cases had either typical or atypical symptoms.

The difference between seroprevalence and cumulative reported incidence might be explained by three components: (i) asymptomatic cases (ii) atypical or pauci-symptomatic cases, or (iii) the lack of access to, and uptake, of testing in different regions and countries. The asymptomatic proportion found in studies of quarantine is around 17% [32], and so would only explain a small proportion of the difference. The reports of symptoms suggest that atypical symptoms, such as anosmia, and as well as fever and cough were common in the seropositive but undetected cases. We further examined the difference between seroprevalence and cumulative incidence by using a cumulative incidence imputed from the COVID-19 death rates. A notable example is the study in North-West Iran where the apparent case fatality rate is amongst the highest in the world, and there is also some evidence of under reporting of COVID-19 deaths based on the comparison of excess deaths.

Strengths of this review lie in the thorough search for published and unpublished literature, strict inclusion criteria, and critical appraisal potential studies. However, there are several limitations. First, while we excluded several studies because of their volunteer and/or responder bias, several of the included studies still had significant degrees of non-response. Second, the accuracy of the serological tests used was often unclear. A particular concern was the specificity and possibility of false positive results in lower prevalence settings leading to potential overestimation of seroprevalence.[6] For example, a specificity of 98% implies a 2% false positive rate even in populations with few past infections. Third, to impute cumulative case incidence we assumed a “true” case fatality rate of 1% for all populations[11] and did not allow for any lag-time in using the mortality data. Finally, the inadequate reporting of many studies, particularly the preprints, made the task of data extraction difficult. Many authors did not respond to data-related questions emailed to the corresponding author.

There has been a couple of previous reviews of seroprevalence studies, but these focused on using the studies to infer the infection fatality rate.[33, 34] We excluded some of the primary studies they included because of the poor sampling methods, with high risk of bias from the involvement of volunteers or low response rates. However, both reviews also demonstrated a substantial variation in the seroprevalence rates but with an even greater range than our review because of the inclusion of studies with high risk of bias. The estimated under-ascertainment of infections based on seroprevalence was 6 to 24 times the number of cumulative reported cases in a study from the United States [35], most of the areas they investigated had an estimated infection rates at least 10 times greater than the reported cases, which was similar to our findings.

The results of this review have several implications for policy and practice. First, in all studies the estimated seroprevalences falls well short of that required for herd immunity suggesting that herd immunity is unlikely to be achieved without mass vaccinations. Additionally, infection fatality rates are shown to increase severalfold as the age of the people advance, further proving that herd immunity should not be pursued through the natural course of a pandemic. [36] Reaching herd immunity does not guarantee low or zero disease prevalence and susceptible individuals will still remain at risk of infection.[37] Second, studies in regions with relatively thorough symptom-based testing and detection show only a modest gap between the seroprevalence and the case cumulative incidence, suggesting that much of the gap between reported cases and seroprevalence is likely to be due to undetected symptomatic cases. Third, the short serial interval, days 3 to 5, post-exposure enables the exposed person to become a source of transmission prior to developing symptoms.[38] Estimating cumulative cases on test-and-trace approaches that test only symptomatic contacts will underestimates of community seroprevalence. Fourth, the variation and incompleteness of methods used by the studies points to the need for better standardisation, design, and reporting of seroprevalence studies, including the need for better questioning and reporting of subjects, prior history of RT-PCR testing, and history of symptoms.

Routine testing for an immune response to COVID-19 in recovered patients allows not only evaluation of the transmissibility of infection in general and specific populations, but would provide improved estimations of attack rates and infection fatality rates, estimates of possible immunity and evidence of reinfection.[39-41] The detection of antibodies established from the studies we analysed does not infer herd immunity levels in their populations. SARS-CoV- 2 shares 79.6% sequence identity to SARS-CoV [42], and the peak level of IgG/neutralising antibodies in recovered SARS-CoV patients occurred at 4-6 months before declining.[43] Knowing the duration of immunity could inform strategic public health approaches until a vaccine is available. Accurate estimates of immunity will not only require repeat antibody testing among the population, but also establishing the association between a positive antibody response and protective immunity against the disease. The current unknown duration of IgG response and its association with disease immunity also raises questions about the validity of an “immunity passport”, especially past a probable peak at 4-6 months post infection.[43, 44]

Findings of this review should help inform policy globally, but also trigger improved research methods and better reporting of any future studies on seroprevalence. When there is a large gap between seroprevalence estimates and incidence rates, strategies to extend case finding and testing needs to be implemented. Evidence-based and targeted public health measures informed by accurate real-world data will help us successfully navigate the uncertain dynamics of this new pandemic.

## Data Availability

The authors declare that data supporting the findings of this study are available within the paper and its supplementary files.

## Authors’ contributions

PG conceived the study and co-designed with OB. JC led the literature searches including backward and forward citation analysis. OB and CCD conducted the parallel title, abstract and full text screening, and risk of bias assessment. OB, CCD, PG, KB, DPR did data extractions, analysis, and summary figures and tables. MLM provided expertise in interpretation of the findings and drafting of the manuscript. All authors contributed to resolving disagreements throughout the study conduct and to writing of the manuscript.

## Acknowledgment

We thank the authors of eligible manuscripts for their replies to our queries.

## Data availability statement

The data underlying this article are available in the article and its online supplementary files.

## Supporting information

**S1. Full search strategy**.

**S2. Table of excluded full text articles with reasons**.

**S3. Full signalling questions for risk of bias assessment**.

**S4. Table with serological test sensitivity and specificity, adjustment details of studies and sources for cumulative incidence data**.

**S5. Figure showing the data collection timeframes of included studies in relation to the rolling 7-day average of confirmed cases in each country**.

