## Supplementary material for "Comparison of seroprevalence of SARS-CoV-2 infections with cumulative and imputed COVID-19 cases: systematic review": S1. Full search strategy.docx

**PubMed**

(("COVID-19"[Supplementary Concept] OR “COVID-19”[tiab] OR COVID19[tiab] OR “COVID 19”[tiab] OR “SARS-CoV-2”[tiab] OR “2019-nCoV”[tiab] OR “Novel coronavirus”[tiab] OR “Coronavirus 2019”[tiab] OR “Coronavirus 19”[tiab] OR “COVID 2019”[tiab] OR "2019 ncov"[tiab] OR “Wuhan coronavirus”[tiab])

AND

("COVID-19 diagnostic testing"[Supplementary Concept] OR Seroprevalence[tiab] OR “Serological test”[tiab] OR “Serological tests”[tiab] OR ((Infection[tiab] OR Detecting[tiab]) and (Antibody[tiab] OR Antibodies[tiab])))

AND

(Letter[pt] OR "Epidemiologic Studies"[Mesh] OR “Case-control studies”[Mesh] OR “Cohort Studies”[Mesh] OR “Case control”[tiab] OR Longitudinal[tiab] OR Prospective[tiab] OR Retrospective[tiab] OR “Cross sectional”[tiab] OR “Cross-Sectional Studies”[Mesh] OR Investigated[tiab] OR Analysis[tiab] OR Data[tiab] OR Cases[tiab] OR Ratio[tiab] OR "statistics and numerical data"[sh] OR "epidemiology"[sh] OR "Seroepidemiologic Studies"[Mesh]))

OR

((“COVID-19”[ti] OR COVID19[ti] OR “COVID 19”[ti] OR “SARS-CoV-2”[ti] OR “2019-nCoV”[ti] OR “Novel coronavirus”[ti] OR “Coronavirus 2019”[ti] OR “Coronavirus 19”[ti] OR “COVID 2019”[ti] OR "2019 ncov"[ti] OR “Wuhan coronavirus”[ti])

AND

(Seroprevalence[ti]))

**Embase**

(('covid 19'/exp OR COVID-19:ti,ab OR COVID19:ti,ab OR "COVID 19":ti,ab OR SARS-CoV-2:ti,ab OR 2019-nCoV:ti,ab OR "Novel coronavirus":ti,ab OR "Coronavirus 2019":ti,ab OR "Coronavirus 19":ti,ab OR "COVID 2019":ti,ab OR "2019 ncov":ti,ab OR "Wuhan coronavirus":ti,ab)

AND

('seroprevalence'/exp OR Seroprevalence:ti,ab OR "Serological test":ti,ab OR "Serological tests":ti,ab OR ((Infection:ti,ab OR Detecting:ti,ab) AND (Antibody:ti,ab OR Antibodies:ti,ab)))

AND

('letter'/exp OR ‘epidemiology'/exp OR 'case control study'/exp OR 'cohort analysis'/exp OR 'seroepidemiology'/exp OR "Case control":ti,ab OR Longitudinal:ti,ab OR Prospective:ti,ab OR Retrospective:ti,ab OR "Cross sectional":ti,ab OR 'cross-sectional study'/exp OR Investigated:ti,ab OR Analysis:ti,ab OR Data:ti,ab OR Cases:ti,ab OR Ratio:ti,ab))

OR

((COVID-19:ti OR COVID19:ti OR "COVID 19":ti OR SARS-CoV-2:ti OR 2019-nCoV:ti OR "Novel coronavirus":ti OR "Coronavirus 2019":ti OR "Coronavirus 19":ti OR "COVID 2019":ti OR "2019 ncov":ti OR "Wuhan coronavirus":ti)

AND

(Seroprevalence:ti))

**Europe PMC**

(“COVID-19” OR COVID19 OR “COVID 19” OR “SARS-CoV-2” OR “2019-nCoV” OR “Novel coronavirus” OR “Coronavirus 2019” OR “Coronavirus 19” OR “COVID 2019” OR "2019 ncov" OR “Wuhan coronavirus”)

AND

("COVID-19 diagnostic testing" OR Seroprevalence OR ((Infection OR Detecting) and (Antibody OR Antibodies)))
