## Supplementary material for "Comparison of seroprevalence of SARS-CoV-2 infections with cumulative and imputed COVID-19 cases: systematic review": S2. Table of excluded studies.docx

### S2. Table of excluded studies following full text screening

| **#** | **Reference** | **Reason for exclusion** |
| --- | --- | --- |
| 1. | Barna, V., et al. (2020). "The diagnostic value of rapid anti IgM and IgG detecting tests in the identification of patients with SARS CoV-2 virus infection." Orvosi Hetilap **161**(20): 807-812. | High risk sampling: patients |
| 2. | Bassett BA. Strict Lower Bound on the COVID-19 Fatality Rate in Overwhelmed Healthcare Systems. medRxiv. 2020:2020.04.22.20076026. | Modelling study |
| 3. | Bendavid, E., et al. (2020). COVID-19 Antibody Seroprevalence in Santa Clara County, California, medRxiv. <https://doi.org/10.1101/2020.04.14.20062463> | High risk sampling: volunteers |
| 4. | Bennett, S. and M. Steyvers (2020). Estimating COVID-19 Antibody Seroprevalence in Santa Clara County, California. A re-analysis of Bendavid et al, medRxiv. <https://www.medrxiv.org/content/10.1101/2020.04.24.20078824v1> | Re-analysis of Bendavid paper. High risk sampling |
| 5. | Bryan, A., et al. (2020). "Performance Characteristics of the Abbott Architect SARS-CoV-2 IgG Assay and Seroprevalence in Boise, Idaho." J Clin Microbiol. 2020 May 7:JCM.00941-20. doi: 10.1128/JCM.00941-20. | Insufficient data for inclusion |
| 6. | Doi, A., et al. (2020). Estimation of seroprevalence of novel coronavirus disease (COVID-19) using preserved serum at an outpatient setting in Kobe, Japan: A cross-sectional study, medRxiv. <https://www.medrxiv.org/content/10.1101/2020.04.26.20079822v2> | High risk sampling: patients/volunteers |
| 7. | Emmenegger M, De Cecco E, Lamparter D, Jacquat RPB, Ebner D, et al. Population-wide evolution of SARS-CoV-2 immunity tracked by a ternary immunoassay. medRxiv 2020.05.31.20118554; doi: https://doi.org/10.1101/2020.05.31.20118554 | High risk sampling: blood donors |
| 8. | Erikstrup, C., et al. (2020). Estimation of SARS-CoV-2 infection fatality rate by real-time antibody screening of blood donors, MedRxiv. <https://www.medrxiv.org/content/10.1101/2020.04.24.20075291v1> | High risk sampling: blood donors |
| 9. | Fontanet, A., et al. (2020). Cluster of COVID-19 in northern France: A retrospective closed cohort study, medRxiv. <https://www.medrxiv.org/content/10.1101/2020.04.18.20071134v1> | High risk sampling: blood donors |
| 10. | Fujita, K., et al. (2020). Quantitative SARS-CoV-2 antibody screening of healthcare workers in the southern part of Kyoto city during the COVID-19 peri-pandemic period, medRxiv. <https://www.medrxiv.org/content/10.1101/2020.05.12.20098962v2> | High risk sampling: volunteers |
| 11. | Gudbjartsson DF, Helgason A, Jonsson H, Magnusson OT, Melsted P, Norddahl GL, et al. Spread of SARS-CoV-2 in the Icelandic Population. N Engl J Med. 2020. | Only looked at confirmed COVID cases |
| 12. | Modi C, Boehm V, Ferraro S, Stein G, Seljak U. Total COVID-19 Mortality in Italy: Excess Mortality and Age Dependence through Time-Series Analysis. medRxiv. 2020:2020.04.15.20067074. | Modelling study |
| 13. | Ng D, Goldgof G, Shy B, Levine A, Balcerek J, Bapat SP, et al. SARS-CoV-2 seroprevalence and neutralizing activity in donor and patient blood from the San Francisco Bay Area. medRxiv 2020.05.19.20107482; doi: https://doi.org/10.1101/2020.05.19.20107482 | Participants include blood donors and hospitalised patients |
| 14. | Reifer J, Hayum N, Heszkel B, Klagsbald I, Streva VA. SARS-CoV-2 IgG Antibody Responses in New York City. medRxiv 2020.05.23.20111427; doi: https://doi.org/10.1101/2020.05.23.20111427 | High risk sampling: patients |
| 15. | Rinaldi G, Paradisi M. An empirical estimate of the infection fatality rate of COVID-19 from the first Italian outbreak. medRxiv. 2020:2020.04.18.20070912. | Modelling study |
| 16. | Roques L, Klein E, Papaix J, Sar A, Soubeyrand S. Using early data to estimate the actual infection fatality ratio from COVID-19 in France. medRxiv. 2020:2020.03.22.20040915. | Modelling study |
| 17. | Rosenberg ES, Tesoriero JM, Rosenthal EM, Chung R, Barranco MA, Styer LM, et al. Cumulative incidence and diagnosis of SARS-CoV-2 infection in New York. medRxiv 2020; doi: https://doi.org/10.1101/2020.05.25.20113050 | Convenience sampling used |
| 18. | Sandri MT, Azzolini E, Torri V, Carloni S, Tedeschi M, Castoldi M, Mantovani A, Rescigno M. IgG serology in health care and administrative staff populations from 7 hospital representative of different exposures to SARS-CoV-2 in Lombardy, Italy. medRxiv doi: <https://doi.org/10.1101/2020.05.24.20111245> | Insufficient data for inclusion |
| 19. | Slot, E., et al. (2020). Herd immunity is not a realistic exit strategy during a COVID-19 outbreak, Research Square. <https://www.researchsquare.com/article/rs-25862/v1> | High risk sampling: blood donors |
| 20. | Steensels, D., et al. (2020). "Hospital-Wide SARS-CoV-2 Antibody Screening in 3056 Staff in a Tertiary Center in Belgium." Jama. | Insufficient data for inclusion |
| 21. | Takita, M., et al. (2020). Preliminary Results of Seroprevalence of SARS-CoV-2 at Community Clinics in Tokyo, medRxiv. <https://www.medrxiv.org/content/10.1101/2020.04.29.20085449v1> | High risk sampling: patients/volunteers |
| 22. | Thompson, C., et al. (2020). Neutralising antibodies to SARS coronavirus 2 in Scottish blood donors - a pilot study of the value of serology to determine population exposure, medRxiv. <https://www.medrxiv.org/content/10.1101/2020.04.13.20060467v1> | High risk sampling: blood donors |
| 23. | To KK, Cheng VC, Cai J-P, Chan K-H, Chen L-L, Wong L-H, et al. Seroprevalence of SARS-CoV-2 in Hong Kong and in residents evacuated from Hubei province, China: a multicohort study. Lancet Microbe 2020 DOI:https://doi.org/10.1016/S26665247(20)30053-7 | High risk sampling: repatriated people |
| 24. | Valenti, L., et al. (2020). SARS-CoV-2 seroprevalence trends in healthy blood donors during the COVID-19 Milan outbreak, medRxiv. <https://www.medrxiv.org/content/10.1101/2020.05.11.20098442v2> | High risk sampling: blood donors |
| 25. | Villa M, Myers JF, Turkheimer F. COVID-19: Recovering estimates of the infected fatality rate during an ongoing pandemic through partial data. medRxiv. 2020:2020.04.10.20060764. | Modelling study |
| 26. | Weekly report of THL serological population study of the coronavirus epidemic. Helsinki, Finland. <https://thl.fi/en/web/thlfi-en/-/thl-publishes-weekly-results-of-population-study-on-coronavirus-antibodies> | Insufficient data for inclusion |
| 27. | Wilson L. SARS-CoV-2, COVID-19, Infection Fatality Rate (IFR) Implied by the Serology, Antibody, Testing in New York City. SSRN. 2020. | Modelling study |
| 28. | Wu, X., et al. (2020). "Serological tests facilitate identification of asymptomatic SARS-CoV-2 infection in Wuhan, China." J Med Virol. <https://onlinelibrary.wiley.com/doi/abs/10.1002/jmv.25904> | High risk sampling: patients |
| 29. | Xu, X., et al. (2020). "Seroprevalence of immunoglobulin M and G antibodies against SARS-CoV-2 in China." Nature Medicine. <https://www.nature.com/articles/s41591-020-0949-6> | High risk sampling: volunteers |
| 30. | Zou, J., et al. (2020). Antibodies to SARS/CoV-2 in arbitrarily-selected Atlanta residents, medRxiv. <https://www.medrxiv.org/content/10.1101/2020.05.01.20087478v1> | High risk sampling: volunteers |
| 31. | First study carried out on herd immunity of the population in the whole territory of Slovenia. In: Slovenia Ro, editor. Slovenia: Republic of Slovenia. | Insufficient data for inclusion |
| 32. | Collective immunity study SARS-COV-2-CZ-Preval: preliminary results. Czech Republic; 2020. | Insufficient data for inclusion |
| 33. | IU, ISDH release preliminary findings about impact of COVID-19 in Indiana. Indiana: Indiana University. | Insufficient data for inclusion |
| 34. | Swedish study <https://www.folkhalsomyndigheten.se/nyheter-och-press/nyhetsarkiv/2020/maj/forsta-resultaten-fran-pagaende-undersokning-av-antikroppar-for-covid-19-virus/> | Insufficient data for inclusion |
| 35. | Sero-epidemiology COVID-19 in Belgium 2020 <https://www.uantwerpen.be/en/research-groups/vaxinfectio/corona-research/seroepidemiology-co/> | High risk sampling: patients |
| 36. | Armann JP, Unrath M, Kirsten C, Lueck C, Dalpke A, Berner R. Anti-SARS-CoV-2 IgG antibodies in adolescent students and their teachers in Saxony, Germany (SchoolCoviDD19): very low seropraevalence and transmission rates. medRxiv; 2020. | High risk sampling: not random sample of population |
| 37. | Bartolini A, Scapaticci M, Bioli M, Lazzarotto T, Re MC, Mancini R. Immunochromatographic assays for COVID-19 epidemiological screening: our experience. medRxiv; 2020. | High risk sampling: patients |
| 38. | Benucci M, Damiani A, Giannasi G, Li Gobbi F, Quartuccio L, Grossi V, et al. Serological tests confirm the low incidence of COVID-19 in chronic rheumatic inflammatory diseases treated with biological DMARD. Ann Rheum Dis. 2020 Jul 6. | Not a primary study (comment to the editor) |
| 39. | Chamie G, Marquez C, Crawford E, Peng J, Petersen M, Schwab D, et al. SARS-CoV-2 Community Transmission During Shelter-in-Place in San Francisco. medRxiv; 2020. | High risk sampling: patients |
| 40. | Chu VT, Freeman-Ponder B, Lindquist S, Spitters C, Kawakami V, Dyal JW, et al. Investigation and Serologic Follow-Up of Contacts of an Early Confirmed Case-Patient with COVID-19, Washington, USA. Emerg Infect Dis. 2020 Aug;26(8):1671-8. | High risk sampling: not random sample of population |
| 41. | Crovetto F, Crispi F, Llurba E, Figueras F, Gomez-Roig MD, Gratacos E. SEROPREVALENCE AND CLINICAL SPECTRUM OF SARS-CoV-2 INFECTION IN THE FIRST VERSUS THIRD TRIMESTER OF PREGNANCY. medRxiv; 2020. | High risk sampling: not random sample of population |
| 42. | Dave M, Poswal L, Bedi V, Regar L, Vijayvargiya R, Sharma M, et al. Study of antibody-based rapid card test in COVID-19 patients admitted in a tertiary care COVID hospital in Southern Rajasthan. Journal, Indian Academy of Clinical Medicine. 2020;21(1-2):7-11. | High risk sampling: patients |
| 43. | De Marinis Y, Sunnerhagen T, Bompada P, Blackberg A, Yang R, Svensson J, et al. Serology assessment of antibody response to SARS-CoV-2 in patients with COVID-19 by rapid IgM/IgG antibody test. medRxiv; 2020. | Diagnostic test validation study, not seroprevalence |
| 44. | Dietrich M, Norton E, Elliott D, Smira A, Rouelle J, Bond N, et al. SARS-CoV-2 Seroprevalence Rates of Children in Louisiana During the State Stay at Home Order. medRxiv; 2020. | High risk sampling: patients |
| 45. | Dimeglio C, Loubes J-M, Miedougé M, Herin F, Soulat J-M, Izopet J. The real seroprevalence of SARS-CoV-2 in France and its consequences for virus dynamics. Research Square; 2020. | Modelling study |
| 46. | Dingens A, Crawford KHD, Adler A, Steele S, Lacombe K, Eguia R, et al. Serological identification of SARS-CoV-2 infections among children visiting a hospital during the initial Seattle outbreak. medRxiv; 2020. | High risk sampling: patients |
| 47. | Dobi A, Frumence E, Lalarizo Rakoto M, Lebeau G, Vagner D, Sandenon Seteyen A-L, et al. Serological surveys in Reunion Island of the first hospitalized patients revealed that long-lived immunoglobulin G antibodies specific against SARS-CoV2 virus are rapidly vanishing in severe cases. medRxiv; 2020. | High risk sampling: patients |
| 48. | Ebinger J, Botwin G, Albert C, Alotaibi M, Arditi M, Berg A, et al. SARS-CoV-2 Seroprevalence in Relation to Timing of Symptoms. medRxiv; 2020. | Secondary analysis of data |
| 49. | Emmenegger M, De Cecco E, Lamparter D, Jacquat R, Ebner D, Schneider M, et al. Early peak and rapid decline of SARS-CoV-2 seroprevalence in a Swiss metropolitan region. medRxiv; 2020. | Diagnostic test validation study, not seroprevalence |
| 50. | fiore jr, centra m, de carlo a, granato m, rosa a, de feo l, et al. FAR AWAY FROM HERD IMMUNITY TO SARS-CoV-2: results from a survey in healthy blood donors in South Eastern Italy. medRxiv; 2020. | High risk sampling: blood donors |
| 51. | Hallal P, Hartwig F, Horta B, Victora G, Silveira M, Struchiner C, et al. Remarkable variability in SARS-CoV-2 antibodies across Brazilian regions: nationwide serological household survey in 27 states. medRxiv; 2020. | Already included |
| 52. | Hallal PC, Horta BL, Barros AJD, Dellagostin OA, Hartwig FP, Pellanda LC, et al. Trends in the prevalence of COVID-19 infection in Rio Grande do Sul, Brazil: repeated serological surveys. Cien Saude Colet. 2020 Jun;25(suppl 1):2395-401. | Duplicate of above, published in portugese |
| 53. | Herzog S, De Bie J, Abrams S, Wouters I, Ekinci E, Patteet L, et al. Seroprevalence of IgG antibodies against SARS coronavirus 2 in Belgium: a prospective cross-sectional nationwide study of residual samples. medRxiv; 2020. | High risk sampling: not random sampling |
| 54. | Kammon A, El-Arabi A, Erhouma E, Mehemed T, Mohamed O. Seroprevalence of antibodies against SARS-CoV-2 among public community and health-care workers in Alzintan City of Libya. medRxiv; 2020. | High risk sampling: hospital outpatients |
| 55. | McDade T, McNally E, Zelikovich A, D'Aquila R, Mustanski B, Miller A, et al. High seroprevalence for SARS-CoV-2 among household members of essential workers detected using a dried blood spot assay. medRxiv; 2020. | High risk sampling: not random sampling |
| 56. | McLaughlin CC, Doll MK, Morrison KT, McLaughlin WL, O'Connor T, Sholukh AM, et al. High Community SARS-CoV-2 Antibody Seroprevalence in a Ski Resort Community, Blaine County, Idaho, US. Preliminary Results. medRxiv. 2020 Jul 21. | High risk sampling: volunteers |
| 57. | Meyers K, Liu L, Lin W-H, Luo Y, Yin M, Wu Y, et al. Antibody Testing Documents the Silent Spread of SARS-CoV-2in New York Prior to the First Reported Case. Research Square; 2020. | Diagnostic test validation study, not seroprevalence |
| 58. | Nopsopon T, Pongpirul K, Chotirosniramit K, Hiransuthikul N. COVID-19 Antibody in Thai Community Hospitals. medRxiv; 2020. | High risk sampling: outpatients |
| 59. | Pancrazzi A, Magliocca P, Lorubbio M, Vaggelli G, Galano A, Mafucci M, et al. Comparison of serologic and molecular SARS-CoV 2 results in a large cohort in Southern Tuscany demonstrates a role for serologic testing to increase diagnostic sensitivity. Clin Biochem. 2020 Jul 21. | High risk sampling: patients |
| 60. | Percivalle E, Cambiè G, Cassaniti I, Nepita EV, Maserati R, Ferrari A, et al. Prevalence of SARS-CoV-2 specific neutralising antibodies in blood donors from the Lodi Red Zone in Lombardy, Italy, as at 06 April 2020. Euro Surveill. 2020 Jun;25(24). | High risk sampling: blood donors |
| 61. | Perez-Saez J, Lauer S, Kaiser L, Regard S, Delaporte E, Guessous I, et al. Serology-informed estimates of SARS-COV-2 infection fatality risk in Geneva, Switzerland. medRxiv; 2020. | Not seroprevalence study |
| 62. | Qin X, Shen J, Dai E, Li H, Tang G, Zhang L, et al. The change pattern and significance of IgM and IgG in the progress of COVID-19 disease. medRxiv; 2020. | High risk sampling: patients |
| 63. | Reifer J, Hayum N, Heszkel B, Klagsbald I, Streva VA. SARS-CoV-2 IgG antibody responses in New York City. Diagn Microbiol Infect Dis. 2020 Jul 21;98(3):115128. | High risk sampling: patients |
| 64. | Rosenberg E, Tesoriero J, Rosenthal E, Chung R, Barranco M, Styer L, et al. Cumulative incidence and diagnosis of SARS-CoV-2 infection in New York. medRxiv; 2020. | High risk sampling: not random sampling |
| 65. | Solbach W, Schiffner J, Backhaus I, Burger D, Staiger R, Tiemer B, et al. Antibody profiling of COVID-19 patients in an urban low-incidence region in Northern Germany. medRxiv; 2020. | High risk sampling: patients |
| 66. | Song SK, Lee DH, Nam JH, Kim KT, Do JS, Kang DW, et al. IgG Seroprevalence of COVID-19 among Individuals without a History of the Coronavirus Disease Infection in Daegu, Korea. J Korean Med Sci. 2020 Jul 27;35(29):e269. | High risk sampling: patients |
| 67. | Stadlbauer D, Tan J, Jiang K, Hernandez M, Fabre S, Amanat F, et al. Seroconversion of a city: Longitudinal monitoring of SARS-CoV-2 seroprevalence in New York City. medRxiv; 2020. | High risk sampling: not random sampling |
| 68. | Takita M, Matsumura T, Yamamoto K, Yamashita E, Hosoda K, Hamaki T, et al. Geographical Profiles of COVID-19 Outbreak in Tokyo: An Analysis of the Primary Care Clinic-Based Point-of-Care Antibody Testing. J Prim Care Community Health. 2020 Jan-Dec;11:2150132720942695. | High risk sampling: volunteers |
| 69. | Zhang C, Lin L, Tang D, Liu F, Li M, Li Q, et al. Antibody responses to SARS-CoV-2 in healthy individuals returning to Shenzhen. J Med Virol. 2020 Jul 25. | High risk sampling: not random sampling |
| 70. | Caban-Martinez AJ, Schaefer-Solle N, Santiago K, Louzado-Feliciano P, Brotons A, Gonzalez M, et al. Epidemiology of SARS-CoV-2 antibodies among firefighters/paramedics of a US fire department: a cross-sectional study. Occup Environ Med. 2020 Aug 6. | Not random population sampling |
| 71. | Cavicchiolo ME, Trevisanuto D, Lolli E, Mardegan V, Saieva AM, Franchin E, et al. Universal screening of high-risk neonates, parents, and staff at a neonatal intensive care unit during the SARS-CoV-2 pandemic. Eur J Pediatr. 2020 Aug 7:1-7. | High risk sampling: patients |
| 72. | Clarke C, Prendecki M, Dhutia A, Ali MA, Sajjad H, Shivakumar O, et al. High Prevalence of Asymptomatic COVID-19 Infection in Hemodialysis Patients Detected Using Serologic Screening. Journal of the American Society of Nephrology : JASN. 2020. | High risk sampling: patients |
| 73. | Cohen D, Marlowe G, Contreras G, Sosa MA, Mendoza JM, Lenz O, et al. Assessment of a Laboratory-Based SARS-CoV-2 Antibody Test Among Hemodialysis Patients: A Quality Improvement Initiative. medRxiv; 2020. | High risk sampling: patients |
| 74. | Crovetto F, Crispi F, Llurba E, Figueras F, Gómez-Roig MD, Gratacós E. Seroprevalence and presentation of SARS-CoV-2 in pregnancy. Lancet. 2020 Aug 6. | High risk sampling: patients |
| 75. | De Vriese AS, Reynders M. IgG Antibody Response to SARS-CoV-2 Infection and Viral RNA Persistence in Patients on Maintenance Hemodialysis. Am J Kidney Dis. 2020 Jun 5. | High risk sampling: patients |
| 76. | Halatoko WA, Konu YR, Gbeasor-Komlanvi FA, Sadio AJ, Tchankoni MK, Komlanvi KS, et al. Prevalence of SARS-CoV-2 among high-risk populations in Lom&eacute (Togo) in 2020. medRxiv; 2020. | Not random population sampling |
| 77. | Lindahl JF, Hoffman T, Esmaeilzadeh M, Olsen B, Winter R, Amer S, et al. High seroprevalence of SARS-CoV-2 in elderly care employees in Sweden. Infection Ecology and Epidemiology. 2020;10(1). | Not random population sampling |
| 78. | Mattern J, Vauloup-Fellous C, Zakaria H, Benachi A, Carrara J, Letourneau A, et al. Post lockdown COVID-19 seroprevalence and circulation at the time of delivery, France. medRxiv; 2020. | Not random population sampling |
| 79. | Njuguna H, Wallace M, Simonson S, Tobolowsky FA, James AE, Bordelon K, et al. Serial Laboratory Testing for SARS-CoV-2 Infection Among Incarcerated and Detained Persons in a Correctional and Detention Facility - Louisiana, April-May 2020. MMWR Morb Mortal Wkly Rep. 2020 Jul 3;69(26):836-40. | Not random population sampling |
| 80. | Payne DC, Smith-Jeffcoat SE, Nowak G, Chukwuma U, Geibe JR, Hawkins RJ, et al. SARS-CoV-2 Infections and Serologic Responses from a Sample of U.S. Navy Service Members - USS Theodore Roosevelt, April 2020. MMWR Morb Mortal Wkly Rep. 2020 Jun 12;69(23):714-21. | Not random population sampling |
| 81. | Tang H, Tian JB, Dong JW, Tang XT, Yan ZY, Zhao YY, et al. Serologic Detection of Latent SARS-CoV-2 Infections in Hemodialysis Centers: A Multi-center, Retrospective Study in Wuhan, China. American journal of kidney diseases : the official journal of the National Kidney Foundation. 2020. | High risk sampling: patients |
| 82. | Torres JP, Piñera C, De La Maza V, Lagomarcino AJ, Simian D, Torres B, et al. SARS-CoV-2 antibody prevalence in blood in a large school community subject to a Covid-19 outbreak: a cross-sectional study. Clin Infect Dis. 2020 Jul 10. | Not random population sampling |
| 83. | Liu T, Wu S, Tao H, Zeng G, Zhou F, Guo F, et al. Prevalence of IgG antibodies to SARS-CoV-2 in Wuhan - implications for the ability to produce long-lasting protective antibodies against SARS-CoV-2. medRxiv; 2020. | High risk sampling: healthcare workers |
| 84. | Nopsopon T, Pongpirul K, Chotirosniramit K, Jakaew W, Kaewwijit C, Kanchana S, et al. Seroprevalence of Hospital Staff in Province with Zero COVID-19 cases. medRxiv; 2020. | High risk sampling: healthcare workers |
| 85. | Asuquo MI, Effa E, Otu A, Ita O, Udoh U, Umoh V, et al. Prevalence of IgG and IgM antibodies to SARS-CoV-2 among clinic staff and patients. medRxiv; 2020. | High risk sampling: healthcare workers |
| 86. | Chibwana MG, Jere KC, Kamng'ona R, Mandolo J, Katunga-Phiri V, Tembo D, et al. High SARS-CoV-2 seroprevalence in Health Care Workers but relatively low numbers of deaths in urban Malawi. medRxiv. 2020 Aug 1. | High risk sampling: healthcare workers |
| 87. | Comar M, Brumat M, Concas MP, Argentini G, Bianco A, Bicego L, et al. COVID-19 experience: first Italian survey on healthcare staff members from a Mother-Child Research hospital using combined molecular and rapid immunoassays test. JAMA. 2020 2020. | High risk sampling: healthcare workers |
| 88. | Flannery D, Gouma S, Dhudasia M, Mukhopadhyay S, Pfeifer M, Woodford E, et al. SARS-CoV-2 Seroprevalence Among Parturient Women. Research Square; 2020. | Not random population sampling |
| 89. | Garcia-Basteiro A, Moncunill G, Tortajada M, Vidal M, Guinovart C, Jimenez A, et al. Seroprevalence of antibodies against SARS-CoV-2 among health care workers in a large Spanish reference hospital. medRxiv; 2020. | High risk sampling: healthcare workers |
| 90. | Jerkovic I, Ljubic T, Basic Z, Kruzic I, Kunac N, Bezic J, et al. SARS-CoV-2 antibody seroprevalence in industry workers in Split-Dalmatia and Sibenik-Knin County, Croatia. medRxiv; 2020. | Not random population sampling |
| 91. | Kraehling V, Kern M, Halwe S, Mueller H, Rohde C, Savini M, et al. Epidemiological study to detect active SARS-CoV-2 infections and seropositive persons in a selected cohort of employees in the Frankfurt am Main metropolitan area. medRxiv. 2020:2020.05.20.20107730. | Not random population sampling |
| 92. | Biggs HM, Harris JB, Breakwell L, Dahlgren FS, Abedi GR, Szablewski CM, et al. Estimated Community Seroprevalence of SARS-CoV-2 Antibodies - Two Georgia Counties, April 28-May 3, 2020. MMWR Morb Mortal Wkly Rep. 2020 Jul 24;69(29):965-70. | Response rate <25% threshold |
| 93. | Brotons C, Serrano J, Fernandez D, Garcia-Ramos C, Ichazo B, Lemaire J, et al. Seroprevalence against COVID-19 and follow-up of suspected cases in primary health care in Spain. medRxiv; 2020. | Not true random sampling, Only asymptomatic people |
| 94. | Feehan AK, Fort D, Garcia-Diaz J, Price-Haywood E, Velasco C, Sapp E, et al. Seroprevalence of SARS-CoV-2 and Infection Fatality Ratio, Orleans and Jefferson Parishes, Louisiana, USA, May 2020. Emerg Infect Dis. 2020 Jul 30;26(11). | Response rate <25% threshold |
| 95. | Gomes CC, Cerutti C, Zandonade E, Maciel ELN, de Alencar FEC, Almada GL, et al. A population-based study of the prevalence of COVID-19 infection in Espirito Santo, Brazil: methodology and results of the first stage. medRxiv; 2020. | not enough info to be includable (can't ascertain cumulative cases for the study population and no response rate info) |
| 96. | Havers FP, Reed C, Lim T, Montgomery JM, Klena JD, Hall AJ, et al. Seroprevalence of Antibodies to SARS-CoV-2 in 10 Sites in the United States, March 23-May 12, 2020. JAMA Intern Med. 2020 Jul 21. | Convenience residual serum analysis from 10 states of USA |
| 97. | Mahajan S, Srinivasan R, Redlich C, Huston S, Anastasio K, Cashman L, et al. Seroprevalence of SARS-CoV-2-Specific IgG Antibodies Among Adults Living in Connecticut Between March 1 and June 1, 2020: Post-Infection Prevalence (PIP) Study. medRxiv; 2020. | Response rate <25% threshold |
| 98. | Menachemi N, Yiannoutsos CT, Dixon BE, Duszynski TJ, Fadel WF, Wools-Kaloustian KK, et al. Population Point Prevalence of SARS-CoV-2 Infection Based on a Statewide Random Sample - Indiana, April 25-29, 2020. MMWR Morb Mortal Wkly Rep. 2020 Jul 24;69(29):960-4. | Response rate <25% threshold |
| 99. | Pagani G, Conti F, Giacomelli A, Bernacchia D, Rondanin R, Prina A, et al. Seroprevalence of SARS-CoV-2 IgG significantly varies with age: results from a mass population screening (SARS-2-SCREEN-CdA). medRxiv; 2020. | Not enough details for inclusion (participant characteristics etc) |
| 100. | Tess B, Granato C, Alves M, Pintao M, Rizzatti E, Nunes M, et al. SARS-CoV-2 seroprevalence in the municipality of Sao Paulo, Brazil, ten weeks after the first reported case. medRxiv; 2020. | not enough info to be includable (can't ascertain cumulative cases for the study population) |
| 101. | Vieira MACS, Vieira CPB, Borba AS, Melo MCC, Oliveira MS, Melo RM, et al. Sequential serological surveys in the early stages of the coronavirus disease epidemic: Limitations and perspectives. Revista da Sociedade Brasileira de Medicina Tropical. 2020;53:1-4. | not enough info to be includable |
| 102. | Laboratory surveillance for SARS-CoV-2 in India: Performance of testing & descriptive epidemiology of detected COVID-19, January 22 - April 30, 2020. Indian J Med Res. 2020 May;151(5):424-37. | High risk sampling: healthcare workers |
| 103. | Amendola A, Tanzi E, Folgori L, Barcellini L, Bianchi S, Gori M, et al. Low seroprevalence of SARS-CoV-2 infection among healthcare workers of the largest children hospital in Milan during the pandemic wave. Infect Control Hosp Epidemiol. 2020 Aug 6:1-6. | High risk sampling: healthcare workers |
| 104. | Augusto J, Menacho K, Andiapen M, Bowles R, Burton M, Welch S, et al. Healthcare Workers Bioresource: Study outline and baseline characteristics of a prospective healthcare worker cohort to study immune protection and pathogenesis in COVID-19. Wellcome Open Res; 2020. | High risk sampling: healthcare workers |
| 105. | Bampoe S, Lucas DN, Neall G, Sceales P, Aggarwal R, Caulfield K, et al. A cross-sectional study of immune seroconversion to SARS-CoV-2 in frontline maternity health professionals. Anaesthesia. 2020 Aug 10. | High risk sampling: healthcare workers |
| 106. | Behrens GMN, Cossmann A, Stankov MV, Witte T, Ernst D, Happle C, et al. Perceived versus proven SARS-CoV-2-specific immune responses in health-care professionals. Infection. 2020;48(4):631-4. | High risk sampling: healthcare workers |
| 107. | Blairon L, Mokrane S, Wilmet A, Dessilly G, Kabamba-Mukadi B, Beukinga I, et al. Large-scale, molecular and serological SARS-CoV-2 screening of healthcare workers in a 4-site public hospital in Belgium after COVID-19 outbreak. J Infect. 2020 Jul 30. | High risk sampling: healthcare workers |
| 108. | Brant-Zawadzki M, Fridman D, Robinson P, Zahn M, German R, Breit M, et al. SARS-CoV-2 antibody prevalence in health care workers: Preliminary report of a single center study. medRxiv; 2020. | High risk sampling: healthcare workers |
| 109. | Bruni M, Cecatiello V, Diaz-Basabe A, Lattanzi G, Mileti E, Monzani S, et al. Persistence of anti-SARS-CoV-2 antibodies in non-hospitalized COVID-19 convalescent health care workers. medRxiv; 2020. | High risk sampling: healthcare workers |
| 110. | Chen Y, Tong X, Wang J, Huang W, Yin S, Huang R, et al. High SARS-CoV-2 antibody prevalence among healthcare workers exposed to COVID-19 patients. J Infect. 2020 Sep;81(3):420-6. | High risk sampling: healthcare workers |
| 111. | Ebinger J, Botwin G, Albert C, Alotaibi M, Arditi M, Berg A, et al. SARS-CoV-2 Seroprevalence Across a Diverse Cohort of Healthcare Workers. medRxiv; 2020 | High risk sampling: healthcare workers |
| 112. | Epstude J, Harsch IA. Seroprevalence of COVID-19 antibodies in the cleaning and oncological staff of a municipal clinic. GMS Hyg Infect Control. 2020;15:Doc18. | High risk sampling: healthcare workers |
| 113. | Eyre D, Lumley S, O'Donnell D, Campbell M, Sims E, Lawson E, et al. Differential occupational risks to healthcare workers from SARS-CoV-2: A prospective observational study. medRxiv; 2020. | High risk sampling: healthcare workers |
| 114. | Favara D, Cooke A, Doffinger R, Houghton S, Budriunaite I, Bossingham S, et al. First results from the UK COVID-19 Serology in Oncology Staff Study (CSOS). medRxiv; 2020. | High risk sampling: healthcare workers |
| 115. | Fernández-Rivas G, Quirant-Sánchez B, González V, Doladé M, Martinez-Caceres E, Piña M, et al. Seroprevalence of SARS-CoV-2 IgG Specific Antibodies among Healthcare Workers in the Northern Metropolitan Area of Barcelona, Spain, after the first pandemic wave. medRxiv; 2020. | High risk sampling: healthcare workers |
| 116. | Galan I, Velasco M, Casas L, Goyanes J, Rodriguez-Caravaca G, Losa J, et al. SARS-CoV-2 SEROPREVALENCE AMONG ALL WORKERS IN A TEACHING HOSPITAL IN SPAIN: UNMASKING THE RISK. medRxiv; 2020. | High risk sampling: healthcare workers |
| 117. | Grant J, Wilmore S, McCann N, Donnelly O, Lai R, Kinsella M, et al. Seroprevalence of SARS-CoV-2 antibodies in healthcare workers at a London NHS Trust. Infect Control Hosp Epidemiol. 2020 Aug 4:1-12. | High risk sampling: healthcare workers |
| 118. | Houlihan C, Vora N, Byrne T, Lewer D, Heaney J, Moore D, et al. SARS-CoV-2 virus and antibodies in front-line Health Care Workers in an acute hospital in London: preliminary results from a longitudinal study. medRxiv; 2020. | High risk sampling: healthcare workers |
| 119. | Hunter BR, Dbeibo L, Weaver C, Beeler C, Saysana M, Zimmerman M, et al. Seroprevalence of SARS-CoV-2 Antibodies Among Healthcare Workers With Differing Levels of COVID-19 Patient Exposure. Infect Control Hosp Epidemiol. 2020 Aug 3:1-7. | High risk sampling: healthcare workers |
| 120. | Iversen K, Bundgaard H, Hasselbalch RB, Kristensen JH, Nielsen PB, Pries-Heje M, et al. Risk of COVID-19 in health-care workers in Denmark: an observational cohort study. Lancet Infect Dis. 2020 Aug 3. | High risk sampling: healthcare workers |
| 121. | Kumar D, Ferreira V, Chruscinski A, Kulasingam V, Pugh T, Dus T, et al. Prospective Observational Study of Screening Asymptomatic Healthcare Workers for SARS-CoV-2 at a Canadian Tertiary Care Center. medRxiv; 2020. | High risk sampling: healthcare workers |
| 122. | Lackermair K, William F, Grzanna N, Lehmann E, Fichtner S, Kucher HB, et al. Infection with SARS-CoV-2 in primary care health care workers assessed by antibody testing. Fam Pract. 2020 Aug 7. | High risk sampling: healthcare workers |
| 123. | Lahner E, Dilaghi E, Prestigiacomo C, Alessio G, Marcellini L, Simmaco M, et al. Prevalence of Sars-Cov-2 Infection in Health Workers (HWs) and Diagnostic Test Performance: The Experience of a Teaching Hospital in Central Italy. Int J Environ Res Public Health. 2020 Jun 19;17(12). | High risk sampling: healthcare workers |
| 124. | Li G, Hu C, He Q, Liu J, Xiong N, Wang H. Apparent and occult infections of medical staff in a COVID-19 designated hospital. J Infect Public Health. 2020 Jul 14. | High risk sampling: healthcare workers |
| 125. | Malickova K, Kratka Z, Luxova S, Bortlik M, Lukas M. Anti-SARS-CoV-2 antibody testing in IBD healthcare professionals: are we currently able to provide COVID-free IBD clinics? Scandinavian Journal of Gastroenterology. 2020:1-3. | High risk sampling: healthcare workers |
| 126. | Martin C, Montesinos I, Dauby N, Gilles C, Dahma H, Van Den Wijngaert S, et al. Dynamic of SARS-CoV-2 RT-PCR positivity and seroprevalence among high-risk health care workers and hospital staff. J Hosp Infect. 2020 Jun 25;106(1):102-6. | High risk sampling: healthcare workers |
| 127. | Morcuende M, Guglielminotti J, Landau R. Anesthesiologists' and intensive care providers' exposure to COVID-19 infection in a New York City academic center: a prospective cohort study assessing symptoms and COVID-19 antibody testing. Anesth Analg. 2020 Jun 9. | High risk sampling: healthcare workers |
| 128. | Nakamura A, Sato R, Ando S, Oana N, Nozaki E, Endo H, et al. Seroprevalence of Antibodies to SARS-CoV-2 in Healthcare Workers in Non-epidemic Region: A Hospital Report in Iwate Prefecture, Japan. medRxiv; 2020. | High risk sampling: healthcare workers |
| 129. | Olalla J, Correa AM, Martín-Escalante MD, Hortas ML, Martín-Sendarrubias MJ, Fuentes V, et al. Search for asymptomatic carriers of SARS-CoV-2 in healthcare workers during the pandemic: a Spanish experience. Qjm. 2020 Aug 10. | High risk sampling: healthcare workers |
| 130. | Pallett SJC, Rayment M, Patel A, Fitzgerald-Smith SAM, Denny SJ, Charani E, et al. Point-of-care serological assays for delayed SARS-CoV-2 case identification among health-care workers in the UK: a prospective multicentre cohort study. Lancet Respir Med. 2020 Jul 24. | High risk sampling: healthcare workers |
| 131. | Plebani M, Padoan A, Fedeli U, Schievano E, Vecchiato E, Lippi G, et al. SARS-CoV-2 serosurvey in Health Care Workers of the Veneto Region. medRxiv; 2020. | High risk sampling: healthcare workers |
| 132. | Poulikakos D, Sinha S, Kalra PA. SARS-CoV-2 antibody screening in healthcare workers in a tertiary centre in North West England. Journal of Clinical Virology. 2020;129. | High risk sampling: healthcare workers |
| 133. | Psichogiou M, Karabinis A, Pavlopoulou I, Basoulis D, Petsios K, Roussos S, et al. Antibodies against SARS-CoV-2 among health care workers in a country with low burden of COVID-19. medRxiv; 2020. | High risk sampling: healthcare workers |
| 134. | Reiter T, Pajenda S, Wagner L, Gaggl M, Atamaniuk J, Holzer B, et al. Covid-19 serology in nephrology health care workers. medRxiv; 2020. | High risk sampling: healthcare workers |
| 135. | Rivett L, Sridhar S, Sparkes D, Routledge M, Jones NK, Forrest S, et al. Screening of healthcare workers for SARS-CoV-2 highlights the role of asymptomatic carriage in COVID-19 transmission. Elife. 2020 May 11;9. | High risk sampling: healthcare workers |
| 136. | Rudberg A-S, Havervall S, Manberg A, Jernbom Falk A, Aguilera K, Ng H, et al. SARS-CoV-2 exposure, symptoms and seroprevalence in health care workers. medRxiv; 2020. | High risk sampling: healthcare workers |
| 137. | Sandri MT, Azzolini E, Torri V, Carloni S, Tedeschi M, Castoldi M, et al. IgG serology in health care and administrative staff populations from 7 hospital representative of different exposures to SARS-CoV-2 in Lombardy, Italy. medRxiv; 2020. | Duplicate |
| 138. | Schmidt SB, Grüter L, Boltzmann M, Rollnik JD. Prevalence of serum IgG antibodies against SARS-CoV-2 among clinic staff. PLoS One. 2020;15(6):e0235417. | High risk sampling: healthcare workers |
| 139. | Sotgiu G, Barassi A, Miozzo M, Saderi L, Piana A, Orfeo N, et al. SARS-CoV-2 specific serological pattern in healthcare workers of an Italian COVID-19 forefront hospital. BMC Pulm Med. 2020 Jul 29;20(1):203. | High risk sampling: healthcare workers |
| 140. | Steensels D, Oris E, Coninx L, Nuyens D, Delforge ML, Vermeersch P, et al. Hospital-Wide SARS-CoV-2 Antibody Screening in 3056 Staff in a Tertiary Center in Belgium. JAMA - Journal of the American Medical Association. 2020;324(2):195-7. | Duplicate |
| 141. | Stubblefield WB, Talbot HK, Feldstein L, Tenforde MW, Rasheed MAU, Mills L, et al. Seroprevalence of SARS-CoV-2 Among Frontline Healthcare Personnel During the First Month of Caring for COVID-19 Patients - Nashville, Tennessee. Clin Infect Dis. 2020 Jul 6. | High risk sampling: healthcare workers |
| 142. | Tu D, Shu J, Wu X, Li H, Xia Z, Zhang Y, et al. Immunological detection of serum antibodies in pediatric medical workers exposed to varying levels of SARS-CoV-2. J Infect. 2020 Jul 25. | High risk sampling: healthcare workers |
| 143. | Vidal-Anzardo M, Solis G, Solari L, Minaya G, Ayala-Quintanilla B, Astete-Cornejo J, et al. Evaluation under field conditions of a rapid test for detection of IgM AND IgG antibodies against SARS-CoV-2. Revista Peruana de Medicina Experimental y Salud Publica. 2020;37(2):203-9. | High risk sampling: healthcare workers |
| 144. | Woon YL, Lee YL, Chong YM, Ayub NA, Krishnabahawan SL, Lau JFW, et al. Serology surveillance of anti-SARS-CoV-2 antibodies among asymptomatic healthcare workers in Malaysian healthcare facilities designated for COVID-19 care. Research Square; 2020. | High risk sampling: healthcare workers |
| 145. | Xiong S, Guo C, Dittmer U, Zheng X, Wang B. The prevalence of antibodies to SARS-CoV-2 in asymptomatic healthcare workers with intensive exposure to COVID-19. medRxiv; 2020. | High risk sampling: healthcare workers |
| 146. | Xu X, Sun J, Nie S, Li H, Kong Y, Liang M, et al. Seroprevalence of immunoglobulin M and G antibodies against SARS-CoV-2 in China. Nat Med. 2020 Aug;26(8):1193-5. | High risk sampling: healthcare workers |
| 147. | Australian Government. DoH. COVID-19, Australia: Epidemiology Report 20 (Fortnightly reporting period ending 5 July 2020). Commun Dis Intell (2018). 2020 Jul 14;44. | No serology data |
| 148. | Pagani G, Bernacchia D, Conti F, Giacomelli A, Rondanin R, Scolari V, et al. Dynamics of the SARS-CoV-2 epidemic in the earliest-affected areas in Italy: 1 Mass screening for SARS-CoV-2 serological positivity (SARS-2-SCREEN). medRxiv; 2020. | Protocol |
| 149. | Majiya H, Aliyu-Paiko M, Balogu VT, Musa DA, Salihu IM, Kawu AA, et al. Seroprevalence of COVID-19 in Niger State. medRxiv; 2020. | Sample not representative |
| 150. | Wells P, Doores K, Couvreur S, Martin Martinez R, Seow J, Graham C, et al. Estimates of the rate of infection and asymptomatic COVID-19 disease in a population sample from SE England. medRxiv; 2020. | Sampling not random or representative |
| 151. | Ling R, Yu Y, He J, Zhang J, Xu S, Sun R, et al. Seroprevalence and epidemiological characteristics of immunoglobulin M and G antibodies against SARS-CoV-2 in asymptomatic people in Wuhan, China. medRxiv; 2020. | High risk sampling: only asymptomatic people |
| 152. | Nisar MI, Ansari N, Amin M, Khalid F, Hotwani A, Rehman N, et al. Serial population based serosurvey of antibodies to SARS-CoV-2 in a low and high transmission area of Karachi, Pakistan. medRxiv; 2020. | High risk sampling: from high transmission area |
| 153. | Skowronski D, Sekirov I, Sabaiduc S, Zou M, Morshed M, Lawrence D, et al. Low SARS-CoV-2 sero-prevalence based on anonymized residual sero-survey before and after first wave measures in British Columbia, Canada, March-May 2020. medRxiv; 2020. | Sampling not random or representative |
