## Supplementary material for "Comparison of seroprevalence of SARS-CoV-2 infections with cumulative and imputed COVID-19 cases: systematic review": S3. Full signalling questions for risk of bias assessment.docx

1. Was the sampling frame a true or close representation of the target population (all at risk of SARS-CoV-2 infection)?

1a. What method of sampling was used? Complete sampling (census), Random selection, Other?

1b. Is there likely to be bias from selection of a study population that does not match the target population (all at risk of the infection)?

2. Was there incomplete ascertainment of immune status in the study population?

2a. Was testing for SARS-CoV-2 antibodies applied to everyone (or a random sample) of the study population? Is the probability of having an antibody test different for people without symptoms/with mild symptoms/with severe symptoms?

2b. Is there likely to be bias from incomplete ascertainment of immune status in the study population?

3. Were acceptable test methods used to identify individuals in the study population for SARS-C-V-2 immunity?

3a. What tests were used to determine SARS-CoV-2 immune status, and is there an unbiased estimate of the diagnostic accuracy for these? Did the choice of tests (e.g. POCT vs lab test) differ depending on whether symptoms were present, and the severity of symptoms?

3b. Is there likely to be bias from inaccurate tests used to define infection status (false positives especially problematic)

4. Was the same diagnostic test used for all subjects?
5. Was the period of data collection appropriate?
