## Supplementary material for "Comparison of seroprevalence of SARS-CoV-2 infections with cumulative and imputed COVID-19 cases: systematic review": S4. Table with sensitivity and specificity, adjustment details and sources for cumulative incidence data.docx

### S4. Table with serological test sensitivity and specificity, adjustment details of studies and sources for cumulative incidence data

| **Study region, country, author** | **Type of serologic test** | **Sensitivity and Specificity** | **Adjustments made or reported** | **Source for cumulative incidence data** |
| --- | --- | --- | --- | --- |
| **Spanish national sero-epidemiological survey**  Pollán et al  Published | IgG and IgM: Orient Gene IgM/IgG, Zhejiang Orient Gene Biotech. | Sensitivity of 88% and 97% for IgM and IgG respectively.  Specificity 100%. | Reported adjustments for sampling weights and post-stratification to allow for differences in non-response rates based on age group, sex, and census-tract income. | <https://ourworldindata.org/> |
| **Brazilian nationwide survey**  Hallal et al  Preprint | IgG and IgM: WONDFO 459 SARS-CoV-2 Antibody Test (Wondfo Biotech Co., Guangzhou, China) | Sensitivity 86.4% and Specificity of 99.6%. | Reported adjustments for sample design and test validity, and for regions. | <https://ourworldindata.org/> |
| **Hungary**  Merkely et al  Published | IgG: SARS-CoV2 IgG Reagent Kit, Abbott Laboratories, Irving, TX, USA. | Sensitivity 100%. Specificity 99.9%. (Independent study by Bryan et al see below). | Reported adjustments for design weights (region, sex, and age categories). | [Study](https://covid19.who.int/region/euro/country/hu) reported |
| **Luxembourg**  Snoeck et al  Preprint | IgG and IgA: CE-labelled ELISA kits most recent versions from Euroimmun. | Sensitivity 92.9% and 85.7%, Specificity 89.2% and 97.8% for IgA and IgG respectively. Combined IgA+G: sens 85.7% spec 99.5% (Study’s own data) | Reported adjustments for design weights (region, sex, and age categories). | Study reported |
| **Rio Grande do Sul, Brazil**  Silviera et al  Published | IgG and IgM: WONDFO SARS-CoV-2 Antibody Test (Wondfo Biotech Co., Guangzhou, China). | From pooling 4 separate validation studies: sensitivity 84.8%, specificity 99%. | Reported adjustments for sample design, population weighting and for sensitivity and specificity. | Rio Grande do Sul <https://covid.saude.gov.br/> |
| **Faroe Island, Denmark**  Petersen et al  Published | IgG and IgM: SARS-CoV-2 Ab ELISA kit (Beijing Wantai Biologic Pharmacy Enterprise) | Sensitivity 94.4%, specificity 100%. | Reported adjustments for sensitivity and specificity using bootstrap methods. | Study reported |
| **LA county, USA**  Sood et al  Published | IgG and IgM: Lateral Flow Immunoassay test (Premier Biotech). | Sensitivity 82.7% and specificity 99.5%. | Reported adjustments for the weighted and unweighted proportion of positive results for accuracy of the test. | <https://github.com/datadesk/california-coronavirus-data/blob/master/latimes-county-totals.csv> |
| **Jersey Island**  The Channel Islands  Report | IgG and IgM: Lateral Flow Immunoassay (Healgen COVID-19 IgG/IgM) | Overall sensitivity:  83.33% Overall specificity: 100%. | Reported adjustment for sensitivity of the test kit. | <https://www.gov.je/Health/Coronavirus/Pages/CoronavirusCases.aspx> |
| **Guilan, Iran**  Shakiba et al  Published | IgG and IgM: VivaDiag COVID‐19 IgM/IgG from VivaChek. | Sensitivity 63.3% for both IgM and IgG. Specificity 100%. | Reported adjustment for population and test characteristics. | Guilan <https://en.wikipedia.org/wiki/COVID-19_pandemic_in_Iran> |
| **Reykjavik, Iceland**  Gudbjartsson et al  Published | pan-Ig: IgM, IgG, & IgA against nucleoprotein (N) (Roche); the receptor binding domain (Wantai); IgM & IgG against N (EDI/Eagle); and IgG & IgA against the spike protein (Euroimmun). | 91.1% represents the lower bound of sensitivity of the combined pan-Ig. | Reported adjusting for age, age squared, sex, and time since qPCR diagnosis. | Study reported |
| **Geneva, Switzerland**  Stringhini et al  Published | IgG: commercially available ELISA for IgG (Euroimmun AG, Lübeck, Germany). | Sensitivity 86.2%, specificity 100%. | Reported adjusting for age and sex. | <https://covid-19-schweiz.bagapps.ch/fr-2.html> |
| **Stockholm, Sweden**  Roxhed et al  Preprint | IgG: commercially available ELISA for IgG against S1 and N proteins | Study determined sensitivity 100%, Specificity 96-100% | Reported: median fluorescence intensity values were log transformed and normalized to adjust for background binding of human IgG. | <https://ourworldindata.org/coronavirus/country/sweden?country=~SWE> |
| **Five university hospital districts, Finland**  Finnish Institute for Health and Welfare Report | IgG: against nucleoprotein and spike glycoprotein S1 and S2, the antigens manufactured by The Native Antigen Company | Study determined Sensitivity: 100%  Specificity: 98% | No adjustment reported or made. | <https://covid19.who.int/region/euro/country/fi> |
| **Gangelt, Germany**  Streeck et al  Published | IgG and IgA: ELISA on the EUROIMMUN Analyzer I platform (most recent CE version for IgG ELISA as of April 2020) | Sensitivity 90.9% Specificity 99.1% | Reported adjusting for age and sex. | North Rhine-Westphalia <https://en.wikipedia.org/wiki/COVID-19_pandemic_in_Germany#North_Rhine-Westphalia> |
| **Barrio Mugica, Buenos Aires, Argentina**  Figar et al  Preprint | IgG: COVIDAR IgG ELISA (Laboratorio Lemos SRL, Buenos Aires, Argentina) | Sensitivity 75% (7 days after symptom onset) and 95% (after 21 days) using RT-PCR as the gold standard. The specificity was 100%. | Reported adjustments for non-response, projection of sample to the entire population, and for age and sex. | Study provided |
| **Utsunomiya, Japan**  Nawa et al  Published | IgG: SARS-CoV2 IgG chemiluminescence assay from Shenzhen YHLO Biotech Co., Ltd., Shenzhen, China | Sensitivity 97.3%, Specificity 96.3% | Reported adjustments to estimate the population-based weighted prevalence using baseline data from the registry, including age, gender, distance to clinic, residential district, and the number of cohabitants of participants and nonparticipants. | <http://www.pref.tochigi.lg.jp/english/documents/hasseijyoukyou0719.pdf> |
| **Neustadt-am-Rennsteig, Germany** Weis et al  Published | IgG: two ELISA:  Epitope Diagnostics Inc., San Diego, USA, Sens: 98.4%, Spec: 99.8%  Euroimmun, Lübeck, Germany: Sens: 90%, Spec: 100%  Four chemiluminescence assays:  Liaison SARS-CoV-2 S1/S2 IgG CLIA, DiaSorin, Saluggia, Italy. Sens: 97.6%, Spec: 99.3%.  Maglumi 2019-nCoV IgG CLIA, Snibe Co., Ltd., Shenzhen, China, Sens: 91.2%, Spec: 100%  Abbott SARS-CoV 2 IgG CMIA, Abbott, Chicago, USA, Sens: 100%, Spec: 99.6%  Elecsys Anti-SARS-CoV-2 ECLIA, Roche, Basel Switzerland) Sens: 100%, Spec: 99.8% | | Study reports adjustment for cluster effects, and age and sex. | Study provided |

Bryan A, Pepper G, Wener MH, Fink SL, Morishima C, Chaudhary A, et al. Performance Characteristics of the Abbott Architect SARS-CoV-2 IgG Assay and Seroprevalence in Boise, Idaho. Journal of Clinical Microbiology. 2020;58(8):e00941-20.
