## Supplementary material for "Comparison of seroprevalence of SARS-CoV-2 infections with cumulative and imputed COVID-19 cases: systematic review": S5. Figure data collection timeframes.docx

### S5. Figure showing the data collection timeframes of included studies in relation to the rolling 7-day average of confirmed cases in each country

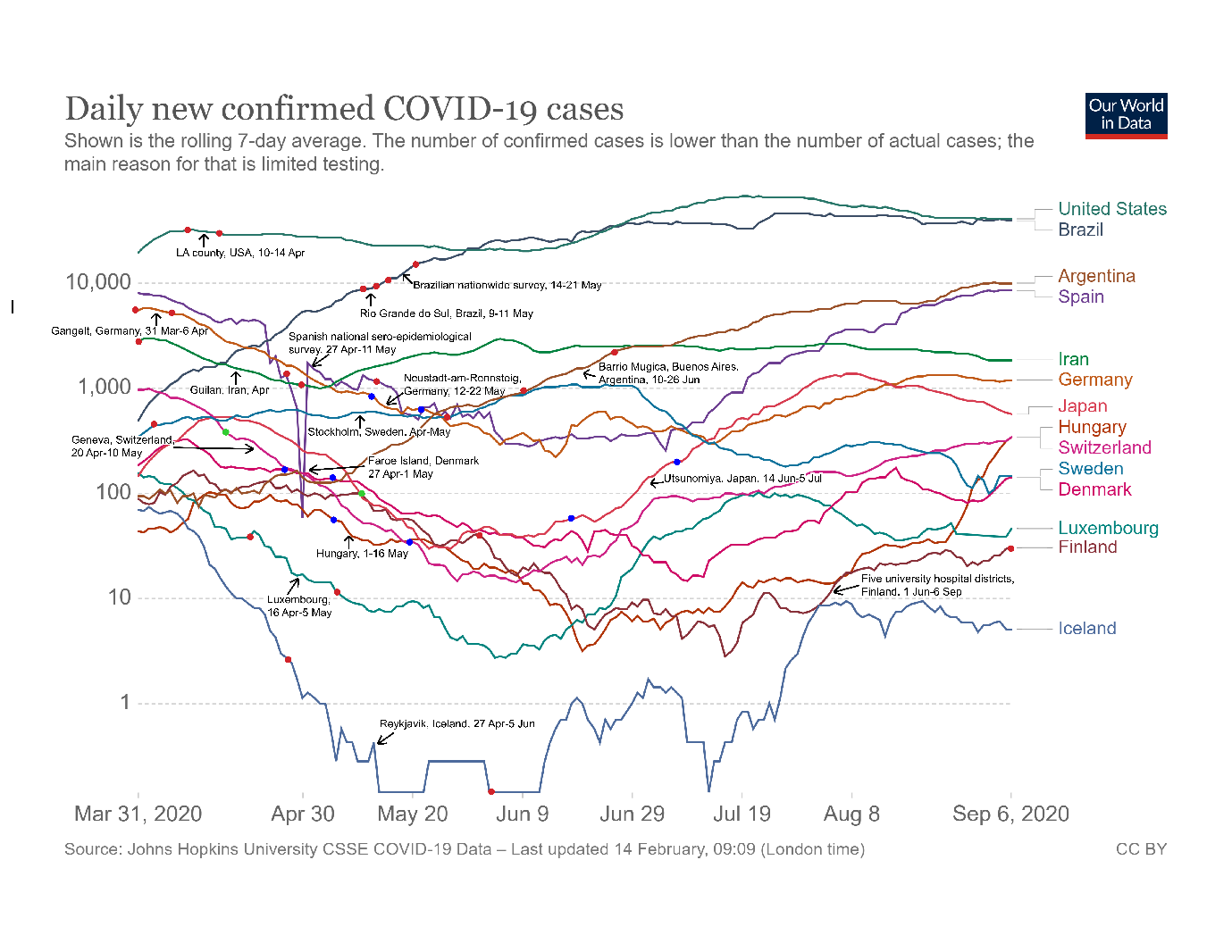

The data collection timeframe for each study is provided in text and marked by two red (or blue if the chart line is reddish) dots on each region’s chart lines.
